# Pathogenesis of Alzheimer’s disease: Involvement of the Choroid Plexus

**DOI:** 10.1101/2021.07.29.21260696

**Authors:** Maria Čarna, Isaac G. Onyango, Stanislav Katina, Dušan Holub, Marketa Nezvedova, Durga Jha, Zuzana Nedelska, Valentina Lacovich, Thijs Vande Vyvere, Ruben Houbrechts, Krystine Garcia-Mansfield, Ritin Sharma, Victoria David-Dirgo, Martin Vyhnalek, Kateřina Texlova, Hernan Chaves, Nadine Bakkar, Lucia Pertierra, Mojmir Vinkler, Hana Markova, Jan Laczo, Kateřina Sheardova, Jan Frič, Antonio Pompeano, Giancarlo Forte, Petr Kaňovsky, Silvie Belaškova, Jiři Damborsky, Jakub Hort, Nicholas T. Seyfried, Robert Bowser, Gustavo Sevlever, Robert A. Rissman, Richard A. Smith, Marian Hajduch, Patrick Pirrotte, Zdeněk Spačil, Eric B. Dammer, Clara Limbäck-Stokin, Gorazd B. Stokin

## Abstract

Aging and Alzheimer’s disease (AD), a major age-related disorder, are both characterized by inflammatory changes in the cerebrospinal fluid (CSF). The origin and the mechanisms underlying these inflammatory changes, however, remain poorly understood. Here, we report that aging elicits inflammatory changes in the CSF that become accentuated uniquely in AD compared to other inflammatory and neurodegenerative disorders. We show that the choroid plexus (ChP), which produces CSF, gains a pro-inflammatory profile, exhibits perturbed metabolism and contributes to the CSF changes observed in AD. We then use MRI imaging to establish a correlation between cognitive decline and increased volume of significantly remodelled ChP in patients with AD, and provide clinical relevance to the identified ChP pathology. These findings collectively suggest that ChP, unable to resolve inflammatory insults efficiently over the lifetime, participates in the inflammation and the pathogenesis of AD.

## Introduction

The choroid plexus (ChP) is a grape-like structure protruding from each of the ventricles of the brain. It is the principal source of cerebrospinal fluid (CSF). Composed of a stroma lined by a tight junction-bound epithelium, the ChP harbours a rich network of fenestrated capillaries of peripheral vascular origin(Ghersi-Egea et al., 2018). This specific architecture of the ChP creates the blood-CSF barrier (BCSFB), which secures optimal composition of the CSF(Damkier et al., 2013) and coordinates the crosstalk of inflammatory signals between the brain and the periphery(Cui et al., 2020; Salvesen et al., 2017; Strominger et al., 2018; Yang et al., 2021). Circulating between the ventricular system and the surface of the brain, the CSF provides nutrients and removes waste playing an essential role in brain homeostasis(Mathew et al., 2016; Myung et al., 2018; Silva-Vargas et al., 2016).

Recent work reveals that the structure and function of the ChP change dramatically during aging(Baruch et al., 2014; Serot et al., 2000; Zhu et al., 2018) and in Alzheimer’s disease (AD), a major age-related disorder(Kant et al., 2018; Serot et al., 2000). These changes are further corroborated in animal models of AD(Brkic et al., 2015; Gonzalez-Marrero et al., 2015), which also show elevated cytokine levels in the ChP(Balusu et al., 2016a; Baruch et al., 2015; Brkic et al., 2015; Steeland et al., 2018) and reversal of select AD phenotypes by attenuating the inflammatory response of the ChP(Baruch et al., 2015; Xu et al., 2020). In agreement with this work, besides impaired production and composition(Gate et al., 2020; Silverberg et al., 2001; Taipa et al., 2019), the CSF demonstrates inflammatory changes involving increased levels of glial markers(Llorens et al., 2017) and cytokines in both aging(Hu et al., 2019; Racine et al., 2019; Sala-Llonch et al., 2017) and AD(Brosseron et al., 2020; Craig-Schapiro et al., 2010; Hu et al., 2019; Rauchmann et al., 2020; Rauchmann et al., 2019; Taipa et al., 2019). These studies, together with the observation that clonally expanded T-cells patrol the CSF in patients with AD (Gate et al., 2020), suggest roles of innate and adaptive immunity in aging and the pathogenesis of AD. Despite established relationships between inflammation, aging and AD, little attention has been given to the possibility that impaired function of the ChP contributes to the pathogenesis of AD. We here investigate age-dependent inflammatory changes of the CSF and test for the involvement of the ChP in the pathogenesis of AD.

## Results

### CSF changes during aging and in AD

Considering that aging represents the most important risk factor of AD and plays an equally important role in inflammation(Onyango et al., 2021), we first investigated the effects of aging on the relative abundance in CSF of proteins in a large, well-established CSF proteomic cohort representative of healthy individuals and patients with AD dementia (Data S1)(Higginbotham et al., 2020; Johnson et al., 2020). We found that half of the most significant CSF protein changes occurring with aging among the unsegregated full cohort of 297 individuals consists of proteins in pathways involving inflammation, including platelet and neutrophil degranulation, complement cascade and innate immunity (Figure 1A). The most significant enriched non-inflammatory pathway alterations with aging included glucose metabolism, post-translational protein modifications, extracellular matrix remodelling and hemostasis. We then compared the CSF protein changes observed in healthy aging with those found when comparing healthy individuals versus patients with AD. In healthy individuals, aging influenced most significantly CSF protein pathways related to the platelet and neutrophil degranulation and innate immunity as well as pathways involved in extracellular matrix remodelling, post-translational protein modifications and hemostasis (Figure 1B). In contrast to healthy individuals, aging across patients with AD was associated with further enrichment of innate immunity and the complement cascade pathways in addition to amyloid fiber formation. These findings indicate that aging coincides with a notable inflammation signature in the CSF, which is exacerbated in AD.

**Figure 1.**
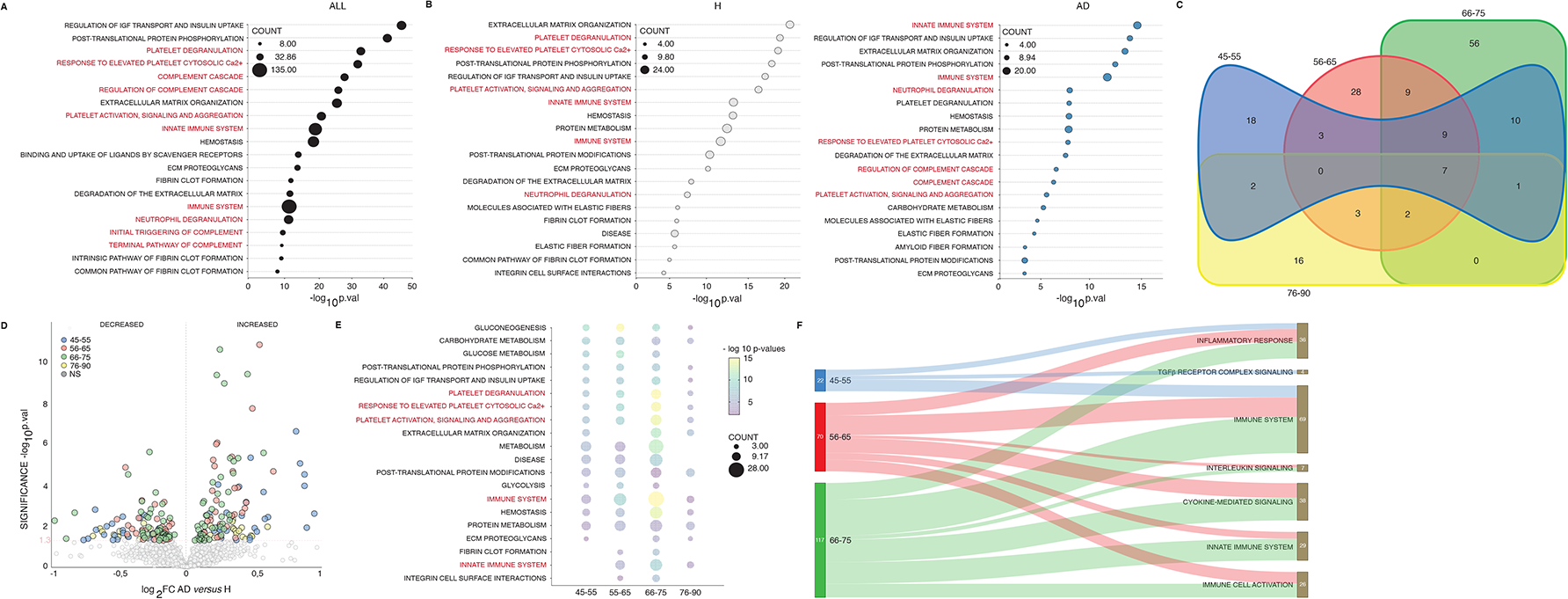
Inflammatory changes in the CSF in aging and AD. (A) The most significant Ingenuity pathways enriched among age-related CSF protein changes in healthy individuals and patients with AD (n=297, p≤0.05, inflammatory changes in red). (B) The most significant Ingenuity pathways enriched among age-related CSF protein changes in healthy individuals (n=147, p≤0.05) and patients with AD (n=150). (C) Venn diagram showing significantly changed CSF proteins in patients with AD compared to healthy individuals in 45-55 (n=29), 56-65 (n=97), 66-75 (n=133) and 76-90 (n=38) year-old age groups (p≤0.05). (D) Volcano plot showing significantly increased and decreased levels of CSF proteins in patients with AD compared to healthy individuals in different age groups (p≤0.05, NS: not significant). (E) Dot plot showing the most significant changes in the Ingenuity pathways enriched in the list of CSF proteins changes in patients with AD compared to healthy individuals in different age groups (p≤0.05). (F) Sankey diagram showing significantly changed inflammation-related biological processes in patients with AD compared to healthy individuals in different age groups (p≤0.05).

### Age-dependent evolution of the inflammatory changes in the CSF of AD patients

To test the association between aging, inflammation and AD further, we re-examined the CSF proteomic cohort of healthy individuals and patients with AD divided into different age groups (Data S2). In all comparisons across the same age groups for healthy individuals versus AD patients, a majority of the significant CSF protein changes consisted of preponderantly increased CSF protein levels (Figure 1C and 1D, Table S1). The most significantly changed pathways in patients with AD compared to healthy individuals in all age groups, and in particular in the 66-75 year-old age group, included platelet degranulation and the immune system in addition to carbohydrate metabolism, post-translational protein modifications, extracellular matrix remodelling and hemostasis (Figure 1E). Intriguingly, the identity and the number of the CSF proteins contributing to the inflammation-related pathway changes in patients with AD compared to healthy individuals differed greatly between the age groups. Patients with AD showed enrichment in the latent transforming growth factor β (TGFβ) binding protein 1 (LTBP1)-driven signaling pathway in the 45-55 year-old age group, extensive changes in the inflammatory response, innate immune system and cytokine-related pathways involving YKL-40, IL6ST, KIT, C1QTNF3 and CD14 in the 56-65 and the 66-75 year-old age groups and the least notable distinction in inflammatory CSF response in the 76-90 year-old age group (Figure 1F). These findings reveal an age-dependent evolution of the inflammatory changes in the CSF of patients with AD compared to healthy individuals.

### Unique inflammatory changes of the CSF in AD

Analysis of the CSF proteome provided compelling evidence that aging plays an important role in the inflammatory changes of the CSF in both healthy people and patients with AD, but it did not interrogate the specificity of the observed inflammatory changes to AD compared to other disorders. To test how specific the observed CSF inflammatory changes are to AD, we compared the CSF protein profile of patients with AD dementia with profiles of patients with mild cognitive impairment due to AD (MCI, negative control), a canonical inflammatory disease, Lyme disease (Lyme, positive control), and a different neurodegenerative disorder, amyotrophic lateral sclerosis (ALS, Figure 2A, Table S2, Data S3). All CSF samples were run on two separate mass spectrometers and only those CSF proteins that were identified by both independent mass spectrometry protocols were included in the analysis (Figure S1). The analysis of the control groups found that patients with MCI showed very few differences, while patients with Lyme exhibited significant CSF protein changes compared to the CSF proteins identified in patients with AD (Figure 2B, Table S3). This finding became most evident during the analysis of the pathways specific to the CSF proteomic profiles of patients with AD versus control disorders. The analysis showed complete overlap in the pathways between patients with AD and MCI (positive control) and significant differences in the pathways between patient with AD and Lyme (positive control) (Figure 2C, Figure S2 and S3). We next compared the CSF from patients with ALS and AD. We found that CSF from patients with ALS exhibited the most significant protein and pathway changes compared to AD patients. Intriguingly, compared to the CSF protein changes identified in AD, the most significant CSF protein changes in ALS as well as in Lyme were found to involve inflammatory pathways playing a role in platelet and neutrophil degranulation, the immune system and the complement cascade. Both ALS as well as Lyme differed from AD in particular, in the inflammatory pathways involving the cytokine and the toll-like receptor cascades (Figure 2D). These findings show that patients with AD demonstrate unique inflammatory changes in the CSF, which differ greatly from the ones observed in inflammatory and other neurodegenerative disorders.

**Figure 2.**
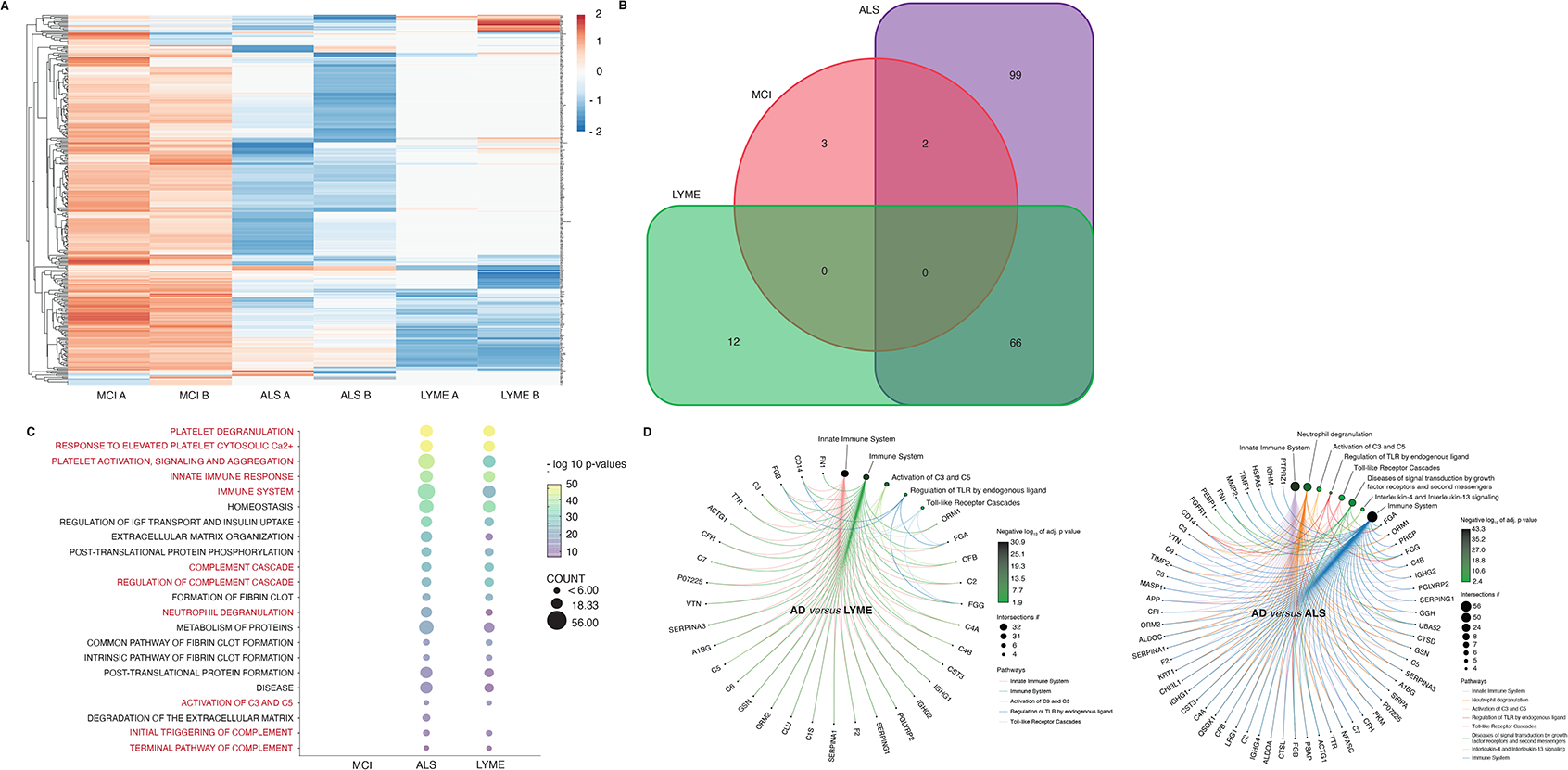
Unique inflammatory changes in the CSF in AD. (A) Heatmap representing hierarchically clustered CSF proteins identified by two independent mass spectrometers and protocols (A and B) in MCI (n=10), AD (n=22), Lyme (n=12) and ALS (n=14) (p≤ 0.05 FDR adjusted t-test). (B) Venn diagram showing significantly changed CSF proteins in MCI, Lyme and ALS in comparison with AD by two independent mass spectrometers and protocols (A and B) (p≤0.05). (C) Dot plot showing the most common significantly changed Reactome-derived pathways in MCI, Lyme and ALS compared to AD (p≤0.05). (D) Circular netplots showing CSF inflammatory proteins and pathways significantly changed in AD compared to Lyme and ALS (p≤0.05).

### Inflammatory changes in the ChP in AD

Inflammatory changes observed in the CSF in patients with AD suggest that ChP, which produces and floats in the CSF, is involved in the transmission of inflammatory signalling in AD. To test whether ChP also exhibits inflammatory signal changes in AD, we first measured cytokines in lysates prepared from frozen post-mortem human ChP by a well-established ELISA array (ELISArray, Data S4). In contrast to healthy individuals, ChP harvested from patients with AD showed significantly increased levels of IL-2, IL-4, tumor necrosis factor α (TNFα) and TGFβ, but not of IL-5, IL-6, IL-10, IL-12, IL-13, IL-17a, interferon γ and the granulocyte-colony stimulating factor (G-CSF, Figure 3A). To confirm this finding, we separated ChP lysates by SDS-PAGE and probed the blots for select cytokines previously examined using the ELISArray. Blots confirmed significantly increased levels of IL-2, TNFα and TGFβ, but not of IL-6 and IL-10, in the ChP obtained from patients with AD compared to healthy individuals (Figure 3B).

**Figure 3.**
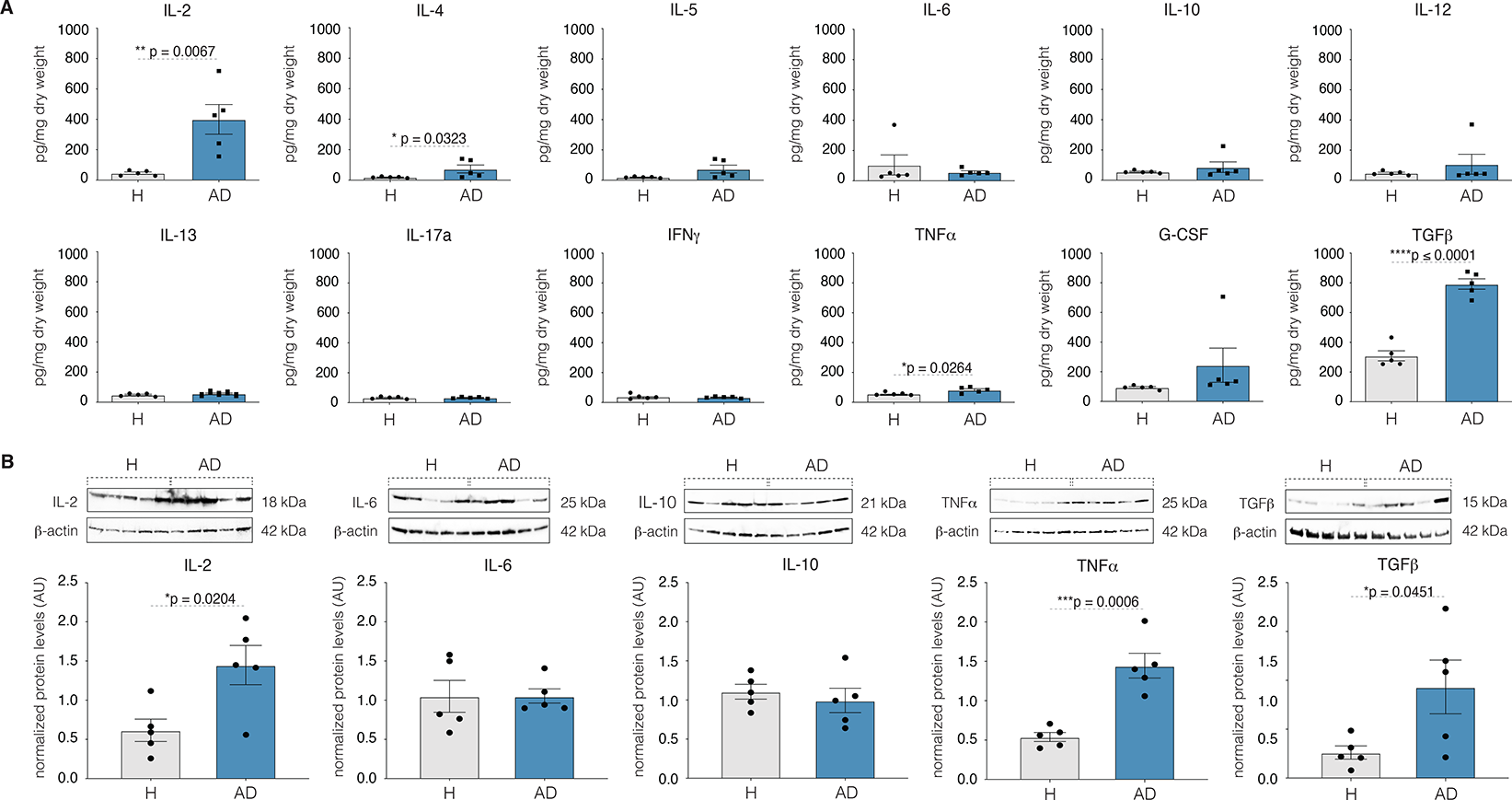
Imbalance between pro- and anti-inflammatory cytokines in the ChP in AD. (A) Levels of IL-2, IL-4, IL-5, IL-6, IL-10, IL-12, IL-13, IL-17a, interferon γ (IFNγ), TNFα, granulocyte colony stimulating factor (G-CSF) and TGFβ in the ChP in H and AD (both n=5). Mean of 3 technical replicates per sample ± s.e.m.; two-sample *t*-test (p≤0.05). (B) Representative Western blots probed for IL-2, IL-6, IL-10, TNFα and TGFβ in the ChP in H and AD (both n=5). All measurements were normalized for β-actin. Mean of 3 technical replicates per sample ± s.e.m.; two-sample *t*-test (p≤0.05).

Since these experiments showed perturbed inflammatory signalling within the ChP harvested from patients with AD, we next examined ChP microscopically for inflammatory and other pathology. Light microscopy examination of haematoxylin and eosin stained, formalin fixed sections of ChP harvested from the lateral cerebral ventricles showed no inflammatory or other overt pathology in patients with AD compared to healthy individuals (Data S5, Figure S4A-S4C). To further test for inflammatory infiltrates in the ChP of patients with AD, we probed ChP sections with anti-CD68 and anti-CD3 antibodies which label macrophages and T lymphocytes, respectively. We found no difference in the number of ChP resident anti-CD68 positive cells between healthy individuals and patients with AD, while the CD3 positive cells were rarely ever observed (Figure S4D-S4F). To exclude region specific pathology, we also examined ChP harvested from the temporal horns of the lateral cerebral ventricles (Data S6). We found no inflammatory infiltrates, AD hallmark lesions or other overt pathology in the ChP obtained from patients with AD compared to healthy individuals (Figure S5). These experiments collectively demonstrate perturbed innate inflammatory signals within the ChP in AD, characterized by a preponderance of pro-inflammatory cytokines.

### Aberrant protein accumulation in the ChP in AD

Considering inflammation invariably alters the function of involved tissues(Nathan, 2002), we next sought to understand whether inflammatory changes identified in the ChP in AD perturb metabolism of the ChP resident proteins secreted to the CSF. To achieve this goal, we randomly selected a subset of proteins from the previously analysed proteomic CSF proteomic dataset and screened whether any protein showing changes in the CSF in patients with AD compared to healthy controls shows similar changes also in the ChP. Out of a subset of 7 randomly selected proteins (ADAM22, APOE, CD44, IGHG2, LDHB, NPC2 and TPI1), we found 4 proteins, CD44, LDHB, NPC2 and TPI1 showing changes in their CSF levels in patients with AD compared to healthy individuals (Figure 4A). We next validated their proteotypic peptide sequences using isotopically labelled standards and measured their concentrations in the ChP by mass spectrometry (Figure S6). We identified 1 out of the 4 proteins, the Niemann-Pick type C2 protein, showing increased levels in AD patient CSF as well as in the ChP of patients with AD compared to healthy individuals. None of the randomly selected proteins with similar CSF levels in healthy individuals and patients with AD showed changes in the ChP (Figure 4B). To confirm the finding of increased NPC2 levels in the ChP of patients with AD, we stained ChP with antibodies against NPC2. Analysis of the scanned NPC2 stained sections showed increased mean intensity of NCP2 immunoreactivity in the ChP harvested from patients with AD compared to healthy individuals (Figure 4C). Confocal imaging of the NPC2-stained sections further showed that physiologically, NPC2 immunoreactivity localizes predominantly to the epithelial subregion of the ChP (Figure 4D). While maintaining this predominantly epithelial distribution of NPC2 immunoreactivity, the ChP harvested from patients with AD demonstrated significantly increased mean intensity of the NPC2 immunoreactivity in the whole as well as in the epithelium, but not in the stromal compartment of the ChP compared to the healthy controls. We also observed that increased mean intensity of the NPC2 immunoreactivity of the ChP acquired a punctate appearance in patients with AD compared to healthy controls. In conclusion, our screening approach identified increased levels of the NPC2 protein in the ChP as well as in the CSF of patients with AD, which suggests impaired metabolism including secretion of the ChP resident proteins in AD. Therefore, the ChP contributes, but is not sufficient to explain all CSF changes observed in AD.

**Figure 4.**
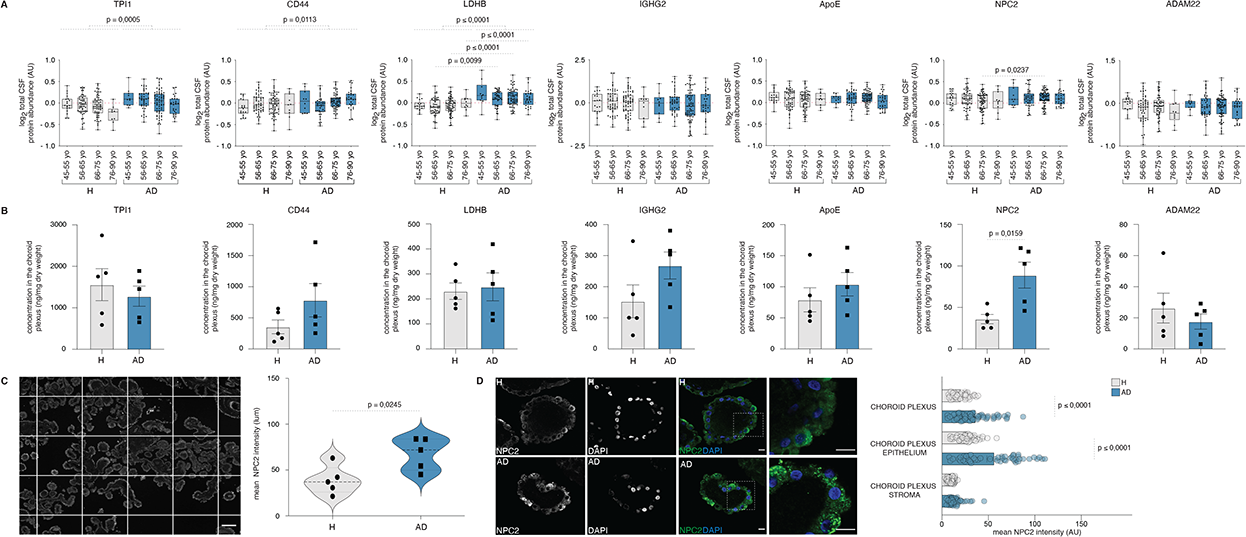
Impaired ChP function in AD. (A) Levels of TPI1, CD44, LDHB, IGHG2, APOE, NPC2 and ADAM22 in the CSF in different age-groups of healthy individuals (n=147) and patients with AD (n=150). (B) Levels of TPI1, CD44, LDHB, IGHG2, APOE, NPC2 and ADAM22 in the ChP of healthy individuals (n=5) and patients with AD (n=5). Mean of 6 technical replicates per sample ± s.e.m.; two-sample *t*-test (p≤0.05). (C) Representative image of a scanned ChP tissue section (magnification bar 100 µ). Mean intensities of NPC2 in the ChP in heathy individuals (n=5) and patients with AD (n=5). Mean of 90 technical replicates per sample ± s.e.m.; two-sample *t*-test (p≤0.05). (D) Mean intensities of NPC2 in the ChP and separately in the ChP epithelium and stroma in healthy individuals (n=5) and patients with AD (n=5) (magnification bar 10 µ). Mean of 3-12 technical replicates ± s.e.m.; repeated measures (p≤0.05).

### Reduced ganglioside levels in the ChP in AD

Considering that previous work linked NPC2 deficiency to lipid metabolism(Peake and Vance, 2010) and increased ganglioside levels(Zhou et al., 2011), we tested whether increased NPC2 levels observed in the ChP harvested from patients with AD also coincides with perturbed lipid metabolism and decreased ganglioside levels. To this end, we systematically measured concentrations of lipids including cholesterol, gangliosides, phosphatidylcholines, phosphatidylethanolamines, sphingomyelins, phosphatidylinositols, phosphatidylserines and sulfatides in the ChP harvested from healthy individuals and patients with AD. The measurements found reduced levels of gangliosides, but not of cholesterol or other classes of lipids in the ChP of patients with AD compared to healthy individuals (Figure 5, Figures S7-S9). Among the gangliosides, we found that levels of GM1, but not of GM2, GM3, GD1a, GD1b, GD2, GD3 and GT1b, decreased in patients with AD compared to healthy individuals. Considering GM1 harbours anti-inflammatory properties(Nikolaeva et al., 2015), the observed reduction in GM1 levels indicates further perturbation of the inflammatory response of the ChP in AD.

**Figure 5.**
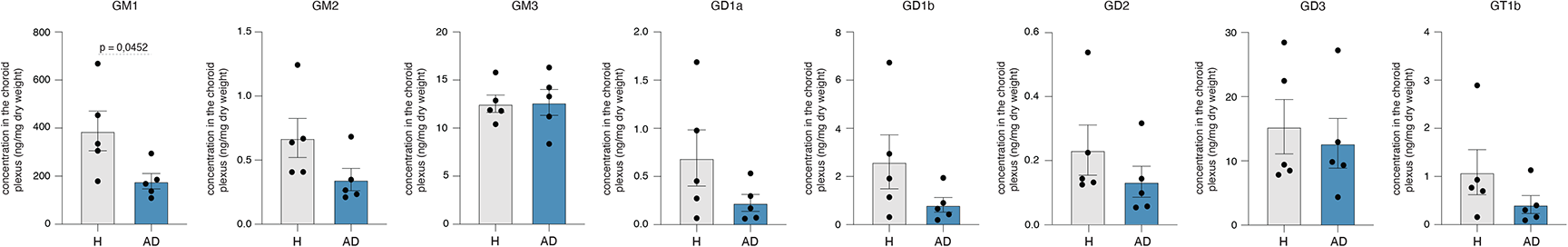
Reduced ganglioside levels in the ChP in AD. (A) Levels of GM1, GM2, GM3, GD1a, GD1b, GD2, GD3 and GT1b gangliosides in the ChP in healthy individuals (n=5) and patients with AD (n=5) ChP. Mean of 6 technical replicates per sample ± s.e.m.; two-sample *t*-test (p≤0.05).

### Enlargement of the ChP in AD

While our findings reveal inflammatory changes and dysfunction of the ChP in AD using tissue samples, the clinical relevance of these findings remains to be critically assessed. To assess their clinical relevance, we imaged brains including ChP in living healthy individuals and patients with AD using magnetic resonance imaging (MRI). We segmented the T1-weighted 3-dimensional (3D) high-resolution MRI brain images acquired using a 1.5 T MRI scanner and measured the volumes of the ChP, the hippocampi (which have been reported to significantly decrease in AD as a positive control)(Seab et al., 1988), and the cerebellar cortices (previously found not to undergo major changes in AD as a negative control)(Zdanovskis, 2021). We observed a significant increase in the ChP volumes of AD patients, while the hippocampal volumes were decreased, and the cerebellar cortical volumes showed no significant changes in patients with AD when compared to healthy individuals (Figure 6A, Data S7). To test this finding further, we created 3-dimensional (3D) representations of the ChP and of the hippocampi (positive control) and re-measured their volumes in a subset of healthy individuals and patients with AD (Data S8). We confirmed a significant increase in the volumes of the ChP, while the hippocampal volumes showed a significant decrease in patients with AD when compared to healthy individuals (Figure 6B). To avoid segmentation-derived measurement errors(Monereo-Sanchez et al., 2021), we next measured ChP volumes in a subset of the acquired MRI brain images manually (Data S9). Manual measurements showed no overlap between the 95% confidence intervals of the ChP volumes in patients with AD compared to healthy individuals, reinforcing the veracity of our finding (Figure 6C). Finally, to confirm the observed enlargement of the ChP in AD further, we acquired MRI brain images using a 3.0 T MRI scanner and measured ChP and hippocampal volumes in a separate cohort of healthy individuals and patients with AD (Data S10). We found a significant increase in the ChP volumes and a significant decrease in the hippocampal volumes (positive control) in patients with AD compared to healthy individuals (Figure S10). To test for clinical relevance of the observed enlargement of the ChP in AD, we probed whether increased ChP volumes correlate with the cognitive performance. We found an inverse correlation between the ChP volumes and the Mini-Mental State Examination (MMSE) scores in patients with AD, but not in healthy individuals (Figure 6D). Therefore, the larger the ChP volume, the poorer the cognitive performance in patients with AD. Taken together, these experiments provide clinical relevance of the inflammatory changes and dysfunction of the ChP found on tissue samples in AD.

**Figure 6.**
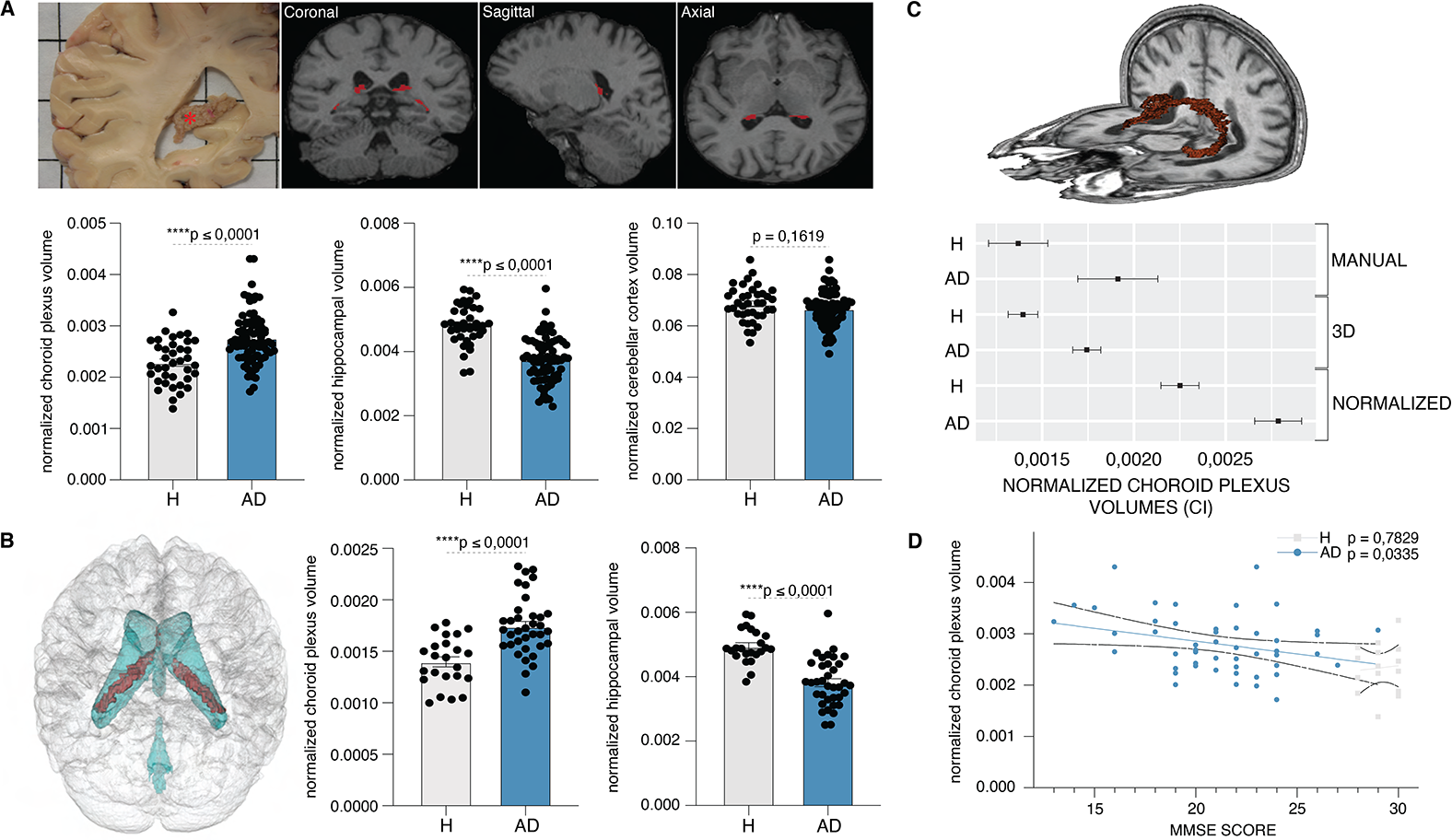
Enlarged ChP in AD. (A) The coronal view of the post-mortem ChP in the right lateral ventricles (red asterisk) and the T1-weighted sequence derived coronal, sagittal and axial views of the ChP (in red). Mean normalized ChP, hippocampal and cerebellar cortical volumes in healthy individuals (n=38) and patients with AD (n=80). Individual measurements ± s.e.m.; two-sample *t*-test (p≤0.05). (B) 3D reconstruction of the brain depicting segmented ChP (red) within the ventricular system (blue). Mean normalized 3D representation-derived ChP and hippocampal volumes in healthy individuals (n=23) and patients with AD (n=36). Single measurements ± s.e.m.; two-sample *t*-test (p≤0.05). (C) Representative 3D image showing manually traced ChP structure (in red). 95% confidence intervals of normalized, 3D representation-derived and manually measured ChP volumes in healthy individuals (n=4) and patients with AD (n=5). (D) Correlation between normalized ChP volumes and MMSE scores in H (n=15) and AD (n=51). Individual measurements ± s.e.m.

### Significant remodelling of the ChP in AD

The observed inflammation, dysfunction and enlargement of the ChP in patients with AD are all indicative of significant structural changes, however, structural changes of the ChP in patients with AD have not yet been investigated. To investigate structural changes of the ChP in patients with AD, we first examined intensities of the T1-weighted 3-D high-resolution MRI ChP images acquired using a 1.5 T MRI scanner in healthy individuals and patients with AD. We found significantly increased ChP intensities in patients with AD compared to healthy individuals (Figure 7A, Data S11). This finding, together with the previously identified enlargement of the ChP in AD, suggested that the ChP undergoes significant structural remodelling in AD. To test whether ChP in AD indeed undergoes significant structural remodelling in AD, we examined changes in the shape of the 3D representations of the ChP and of the hippocampi (as a positive control) in healthy individuals and patients with AD. We found significant atrophy along the anterior margin and hypertrophy in the posterior parts of the ChP in patients with AD compared to healthy individuals, while hippocampi demonstrated previously described atrophic changes across their entire structure(Figure 7B)(Teipel et al., 2006). These experiments demonstrate that ChP undergoes significant structural changes and remodelling in patients with AD.

**Figure 7.**
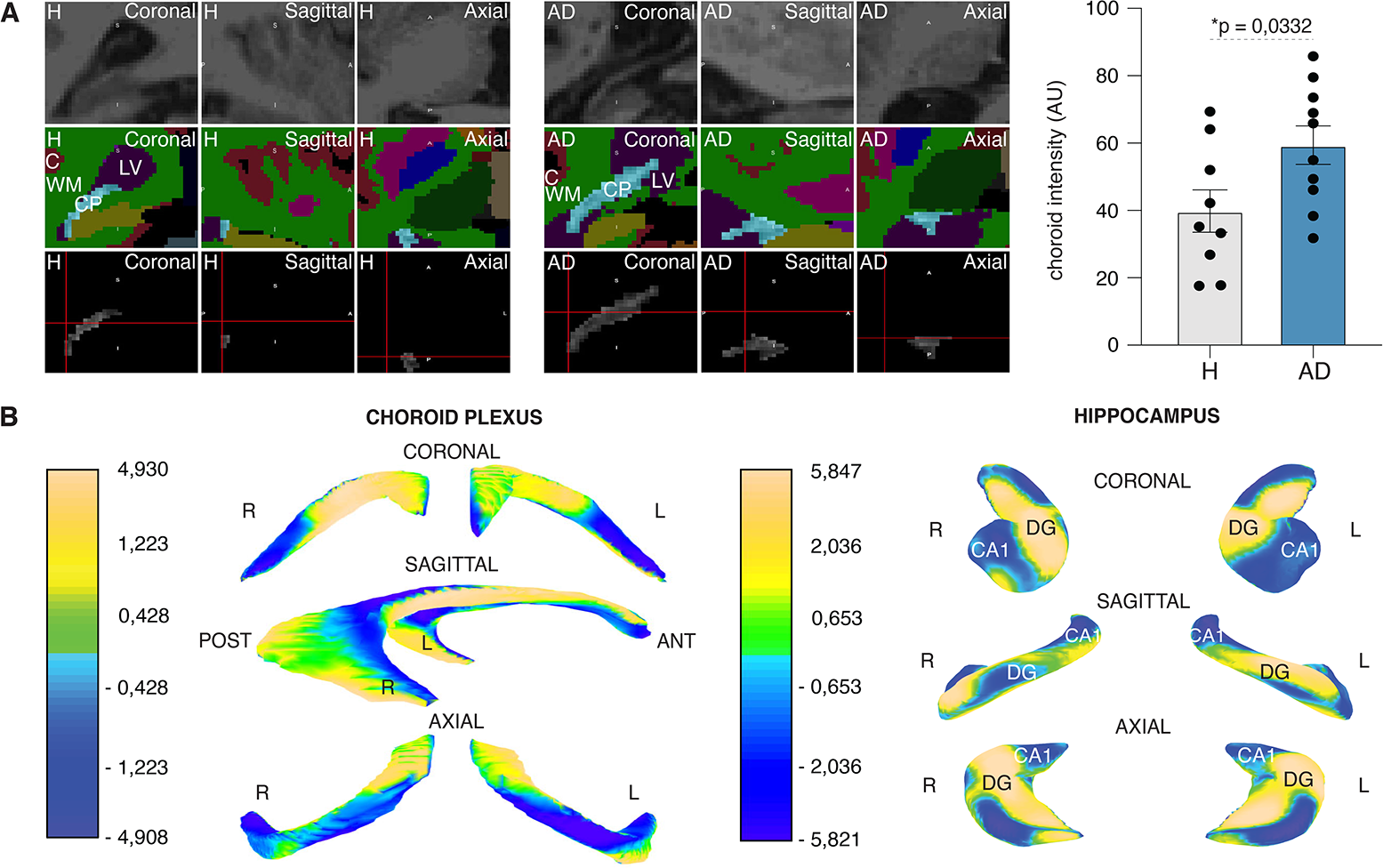
Remodelling of the ChP in AD. (A) Coronal, sagittal and axial views of ChP representative of healthy individuals and patients with AD visualized using T1-weighted sequence (top), FreeSurfer brain structure map (middle) and following ChP segmentation (bottom). Measurements of normalized ChP intensities in healthy individuals (n=9) and patients with AD (n=10). Individual measurements ± s.e.m.; two-sample *t*-test (p≤0.05). (B) Mean changes in the shape of the 3D representations of the ChP and hippocampi in patients with AD (n=36) compared to the healthy individuals (n=23). Single measurements ± s.e.m.; two-sample *t*-test (p≤0.05).

## Discussion

Aging promotes a pro-inflammatory state by disrupting the peripheral immune system and enhancing innate immune system activity with the release of excess pro-inflammatory cytokines and a reduction of anti-inflammatory molecules(Golomb et al., 2020; Gomez et al., 2008; Gruver et al., 2007; Lee et al., 2000; Mrdjen et al., 2018). These changes arising during aging were also recently described in the CSF(Hu et al., 2019; Racine et al., 2019; Sala-Llonch et al., 2017). We here show that inflammatory changes in the CSF in AD represent an accentuated process of aging. We further show that ChP also acquires an enhanced inflammatory state in AD and demonstrates impaired protein and lipid metabolism. Considering the critical role played by the ChP in the homeostasis of the brain, our description here of ChP dysfunction indicates its likely involvement in the pathogenesis of AD.

Our findings are consistent with recent reports noting inflammatory changes in the CSF during aging(Higginbotham et al., 2020; Racine et al., 2019; Sala-Llonch et al., 2017) and in AD(Brosseron et al., 2020; Craig-Schapiro et al., 2010; Hampel et al., 1999; Rangaraju et al., 2018; Rauchmann et al., 2020; Taipa et al., 2019). In contrast to previous reports, we show that the same pathways are activated in the CSF during aging and in AD, except that specific inflammatory pathways such as the innate immune system and the complement cascade are enriched in AD compared to aging. This observation, combined with our and other’s findings of significant differences in the CSF inflammatory pathways between AD and other disorders(Angel et al., 2012; Collins et al., 2015; Costa et al., 2021; Thompson et al., 2018), suggests that the inflammatory changes of the CSF in AD are largely unique to AD and reflect an additional layer of inflammatory changes in addition to an accentuated process of aging. Whether these inflammatory changes exert a protective role in response to aging and/or AD, whether they reflect changes downstream of brain pathology; or whether they otherwise contribute to the pathogenesis of AD awaits further experimental investigation(Albrecht et al., 2021; Bettcher et al., 2018; Craig-Schapiro et al., 2010; Taipa et al., 2019).

We here document inflammatory changes and perturbed protein and lipid metabolism in the ChP harvested from patients with AD compared to healthy controls. These findings are in agreement with the reported upregulation of inflammatory networks in the ChP in patients with AD(Kant et al., 2018; Steeland et al., 2018) and in animal models of AD(Brkic et al., 2015; Mesquita et al., 2015; Xu et al., 2020). Coupled to the observed deleterious effects of the inflammatory molecules on the ChP in culture(Schwerk et al., 2010), these reports suggest that inflammatory changes compromise the structural integrity of the ChP(Brkic et al., 2015; Gonzalez-Marrero et al., 2015; Kant et al., 2018; Pearson et al., 2020; Serot et al., 2000) and contribute to impaired CSF production in AD(Silverberg et al., 2001). Here described pathology discloses impaired function of the ChP, which contributes to the CSF changes(Taipa et al., 2019; Tarkowski et al., 2003), suggests impaired brain homeostasis and a role of ChP in the pathogenesis of AD. Considering disruption of the blood brain barrier (BBB) in AD(Montagne et al., 2015; Sweeney et al., 2018), our findings also raise the question of whether both brain barriers, BBB and BCSFB, fail during aging and play a role in the pathogenesis of AD. A more nuanced understanding of changes in ChP transcytosis-mediated relays across these barriers in AD is also called for(Fung et al., 2018).

During peripheral inflammation, the ChP senses and transmits the information about the peripheral insult to the brain(Balusu et al., 2016b; Baruch et al., 2015; Carloni et al., 2021; Cui et al., 2020; Marques et al., 2009; Salvesen et al., 2017; Strominger et al., 2018; Xu et al., 2020; Yang et al., 2021). During aging, however, despite upregulating its host-defense program(Dani et al., 2021), the ChP undergoes significant involution, which hampers efficient coordination of the inflammatory signals between the brain and the periphery(Baruch et al., 2014; Zhu et al., 2018). Our experiments build on these studies and demonstrate an inflammatory state of the ChP in AD. These findings suggest that at some point during aging, the ChP in AD becomes unable to resolve peripheral inflammatory insults. Since unresolved inflammation is a major driver of disease(Nathan and Ding, 2010), these findings suggest that the ChP could contribute to the pathogenesis of AD also by feeding the brain with continuous “fake” inflammatory signals from the periphery. One can envision that such signals misconstruing peripheral inflammatory status promote a pro-inflammatory state of the brain beyond normal aging observed in AD(Heneka et al., 2018; Hur et al., 2020; Lee et al., 2000; Lu et al., 2004; Wightman et al., 2021).

Finally, we have shown that ChP undergoes significant structural changes and remodelling in patients with AD. These changes are clinically measurable, correlate inversely with cognitive performance of individuals with AD and significantly extend previous work showing a relationship between CSF biomarkers and the ChP volume in AD(Tadayon et al., 2020). Considering the lack of structural studies and, paucity of the volumetric studies of the ChP, it will be imperative to discern the diagnostic and prognostic value of the quantitative and qualitative ChP changes in patients with AD as well as in other neurodegenerative disorders. Considering the paucity of treatments for AD, our work and the work of others suggests that strategies targeting the ChP may be a novel and welcome therapeutic option in the instance of AD and other neurodegenerative disorders(Benson et al., 2010; Smith et al., 2015).

## RESOURCE AVAILABILITY

### Lead contact

Further information and request for resources and reagents should be directed to and will be fulfilled by the lead contact, Gorazd B. Stokin.

### Materials availability

This study did not generate any new unique reagents.

### Data and code availability

All data reported in this paper will be shared by the lead contact upon request. This paper does not report original codes. Any additional information required to reanalyse the data reported in this manuscript is available from the lead contact upon request.

## EXPERIMENTAL MODEL AND SUBJECT DETAILS

### Study participants

Majority of MRI images and CSF samples were acquired through the International Clinical Research Centre of St. Anne’s University Hospital, which funds the Czech Brain Aging Study(Sheardova et al., 2019) running in the Departments of Neurology at Motol and St. Anne’s University Hospitals, Prague and Brno, respectively. A total of 118 living individuals were examined in the original MRI study including 38 cognitively healthy individuals and 80 patients with AD (Supplementary Table 8). The average age of cognitively healthy individuals was 70.0±6.4 years (mean±s.d.) and 75.0±7.6 years for patients with AD. The average MMSE scores for cognitively healthy individuals and patients with AD were 29.0±1.2 and 21.0±3.2, respectively. Randomly selected subsets of individuals of the original MRI study were also used for the assessment of the MRI intensities (Supplementary Table 7), 3D reconstructions (Supplementary Table 9) and for the manual measurements of the ChP (Supplementary Table 10).

Additional 23 living individuals from the Argentina Alzheimer’s Disease Neuroimaging Initiative(Russo et al., 2014) were examined in the confirmatory MRI study including 13 cognitively healthy individuals and 10 patients with AD (Supplementary Table 11).

To maximize the use of publicly available data, we evaluated age-related CSF protein changes using a well-established CSF proteome database (https://www.synapse.org/Consensus). The set consisted in data from a total of 297 individuals, including 147 healthy individuals and 150 patients with AD(Higginbotham et al., 2020; Johnson et al., 2020). The average age of cognitively healthy individuals was 65.1±8.2 years and 68.2±8.3 years for patients with AD (Supplementary Table 4). The average Montreal Cognitive Assessment (MoCA) scores for cognitively healthy individuals and patients with AD were 25.5±3.1 and 14.8±7.4, respectively. A total of 58 living individuals, part of the Czech Brain Aging Study(Sheardova et al., 2019), were evaluated for changes in the CSF including 10 patients with MCI due to AD, 22 patients with AD dementia, 14 patients with ALS(Saul et al., 2020) (all from the Barrow Neurological Institute, Phoenix) and 12 patients with Lyme disease (Supplementary Table 5). The average age was 73.2±7.0, 71.6±9.0, 55.8±8.7 and 69.7±4.3 years for patients with MCI, AD, ALS and Lyme disease, respectively. The average MMSE scores were 26.6±1.4 and 18.3±4.5 for patients with MCI and AD, respectively.

A total of 10 individuals provided post-mortem frozen ChP tissue through the Shiley-Marcos Alzheimer’s Disease Research Centre (ADRC) of the University of California San Diego (UCSD). Tissue included ChP from 5 cognitively healthy individuals and 5 patients with advanced AD (Supplementary Table 1). The average age was 88.0±4.3 years for cognitively healthy individuals and patients with AD. The average MMSE scores were 26.4±3.6 and 19.8±5.0 for cognitively healthy individuals and patients with AD, respectively.

A total of 20 individuals provided post-mortem formalin-fixed paraffin embedded (FFPE) ChP tissue through the Shiley-Marcos UCSD ADRC, including 10 cognitively healthy individuals and 10 patients with advanced AD (Supplementary Table 2). The average age was 84.1±10.7 years for cognitively healthy individuals and 84.1±10.4 years for patients with AD. The average MMSE scores were 28.3±2.8 and 15.9±8.5 for cognitively healthy individuals and patients with AD, respectively.

A total of 44 individuals who are part of the Imperial College Parkinson’s UK Brain Bank(Ruffmann et al., 2016) provided post-mortem stained FFPE ChP slides including 8 healthy individuals, 11 patients with early AD (eAD), 13 patients with late AD (lAD) and 12 patients with both AD and PD (AD/PD) (Supplementary Table 3). The average age was 73.6±9.1, 78.5±11.2, 83.2±9.8 and 79.9±6.7 years for healthy individuals and patients with eAD, lAD and AD/PD, respectively.

Collection of all samples was approved by the Institutional Review Boards of each participating institution with written consent obtained from all individuals. Research was conducted in accordance with the provisions of the Helsinki Declaration.

### Tissue collection

CSF samples were obtained by lumbar puncture with collection and storage of the samples carried out according to a well-established internationally recognized consensus protocol(Saul et al., 2020; Sheardova et al., 2019; Vanderstichele et al., 2012). CSF samples found to be diagnostically unclear or contaminated with blood were excluded from the study. Frozen and formalin fixed paraffin embedded ChP tissue was collected and stored following best practice protocols set forth by the NIH ADRCs. Additional FFPE immune-stained ChP tissue was collected by the Imperial College Parkinson’s UK Brain Bank’s following standardized protocols(Ruffmann et al., 2016). Photographs of post-mortem anatomy of ChP were taken at the Imperial College Parkinson’s UK Brain Bank.

### Cognitive testing

All study individuals underwent regular comprehensive diagnostic assessment, which consisted in extensive clinical evaluation, blood laboratory tests and brain imaging according to NIH and other best practice guidelines(Saul et al., 2020; Sheardova et al., 2019). Clinical evaluation included neurological examination and a battery of neuropsychological tests fulfilling UDS standards. MMSE and the MoCA were used to screen study individuals for cognitive impairment consistent with MCI and dementia. Examinations were performed by board-certified neurologists and clinical psychologists with extensive training and experience in behavioral and cognitive disorders. Clinical phenotypes of MCI and dementia due to AD, PD and ALS were established based on current internationally recognized diagnostic criteria. Lyme disease was diagnosed when individuals demonstrated symptoms and signs of acute polyradiculoneuritis, increased CSF blood count and positive serology for *Borrelia burgdorferi*.

## METHOD DETAILS

### Pathology

Frozen and FFPE ChP samples from the UCSD ADRC were scored based on the Braak and Braak staging of the AD neuropathological changes. Frozen ChP tissue corresponded to Braak and Braak AD scores of 2.0±0 and 5.8±0.4 in cognitively healthy individuals and patients with AD, respectively. FFPE ChP tissue corresponded to Braak and Braak AD scores of 1.3±0.8 and 5.7±0.8 in healthy individuals and patients with AD, respectively. Samples from Imperial College Parkinson’s UK Brain Bank were scored for AD (AD), PD (PD) and mixed AD and PD (AD/PD) neuropathology as previously described(Prokopenko et al., 2019). Brain pathology of healthy individuals (H) and patients with eAD, lAD and AD/PD corresponded to Braak and Braak AD stages of 0.6±0.9, 2.6±0.7, 4.5±0.8 and 4.3±1.2 and to Braak PD stages of 0±0, 0.2±0.4, 1.1±1.5 and 5.5±0.5, respectively. Senile plaques were probed using the primary antibody recognizing amyloid-β residues 17 to 24 (4G8, Covance), neurofibrillary changes using an antibody against phosphorylated tau (AT8, Thermo Fisher Scientific) and Lewy pathology using the α-synuclein clone 42 antibody (#610787, BD Transduction Labs). Neurogenerative pathology was assessed by a board-certified pathologist with training and extensive experience in neurodegenerative disorders according to current neuropathological diagnostic guidelines.

### ELISArray

ChP were homogenized using 0.5 mm glass beads in a Bullet Blender (Next Advance) with two volumes of cold homogenization buffer (20 mM HEPES, 150 mM NaCl, 5 mM EDTA, pH 7.0) supplemented with Halt protease and phosphatase inhibitor cocktail (Thermo Fisher). Samples were then sonicated, centrifuged at 17,000 g for 20 min at 4°C and soluble fractions assessed for their protein content using Pierce BCA protein assay (Thermo Fisher Scientific) and then stored at -80°C. Cytokine levels were measured using Multi-Analyte ELISArray using manufacturer’s instructions (QIAGEN). In brief, absorbance of the samples was measured at 450 nm in the Multiskan Go microplate reader (Thermo Fisher Scientific). Concentration of cytokines was determined based on the absorbance values of negative (buffer) and positive controls (standards for all 12 cytokines examined). All measurements were normalized to the total protein content.

### Western blots

Total amount of 40 µg of protein per homogenated sample (prepared as for the ELISArray measurements) were separated on 4-20% SDS-polyacrylamide gels and transferred to PVDF membranes (Bio-Rad Laboratories). After blocking in 5% BSA in TBS-T, membranes were incubated overnight with primary antibodies against IL-2 (D7A5, Cell Signaling Technology), IL-6 (#6672, Abcam), IL-10 (#34843, Abcam), TNFα (#9739, Abcam) and TGFβ1 (#3711, Cell Signaling Technology) at 4°C. After washing, the membranes were treated with horseradish peroxidase HRP-conjugated secondary antibodies (sc-2357, Santa Cruz Biotechnology) for 1 h at room temperature. Blots were then developed using enhanced chemiluminescence reagent (Clarity Western ECL substrate, Bio-Rad Laboratories Inc) and visualized using ChemiDoc™ MP Imaging System with Image Lab Software (Bio-Rad). All samples were run in single blots for each cytokine. Cytokine levels were normalized to β-actin.

### Immunohistochemistry

FFPE ChP sections were rehydrated with Tissue Clear followed by a 100%, 80% and 50% ethanol to water gradient. Apart from APOE, all section were then subjected to antigen retrieval with citrate buffer of pH 6 at 95°C for 20 (NPC2 and TTR) or 30 (CD3) min or with a Tris-based buffer antigen unmasking solution (Vector Labs) at 95°C for 35 min (CD68). Whenever necessary (NPC2, APOE and TTR), the autofluorescence was quenched using TrueBlack Lipofuscin (Biotium). Sections were blocked either for 10 min with BLOXALL blocking solution (Vector Labs) and 2.5% horse serum (CD3) or for 30 min with 2.5% horse serum (CD68) or 10% with donkey serum (NPC2, APOE and TTR) and then incubated overnight with primary antibodies against NPC2 (1:50, sc-166449, Santa Cruz), APOE (1:100, 178479, Sigma), TTR (1:500, A0002, DAKO), CD3 (1:500, NCL-LCD3-565, Novocastra) or CD68 (1:100, ab955, Abcam). Sections for immunohistochemistry were processed with ImmPRESS Universal Polymer horse anti-mouse/rabbit IgG reagent (Vector Labs) for 30 min, incubated either with NovaRED (CD3, Vector Labs) or ImmPACT DAB EqV solution (CD68, Vector Labs), co-stained with Gill’s hematoxylin (Sigma) and last dehydrated through a 50%, 70% and 100% water to ethanol gradient followed by Tissue Clear and embedment in Eukitt (Sigma). Haematoxylin eosin-stained ChP sections were provided directly by the UCSD ADRC. Sections for immunofluorescence were incubated with goat anti-mouse 647 (NPC2, 1:500, Life Technologies), donkey anti-goat 546 (APOE, 1:500, Life Technologies) or goat anti-rabbit 546 antibodies (TTR, 1:500, Life Technologies) for 1 h at room temperature, co-stained with DAPI for 20 min and mounted using Mowiol (Sigma). Sections stained with secondary antibodies only were used as negative controls.

### Microscopy and image processing

Stained at FFPE ChP sections were imaged using AxioScan.Z1 scanner microscope (Zeiss) at a magnification of 10×. Acquired images were processed and analyzed using the Image Pro Premier 3D software (Media Cybernetics). To examine the distribution of NPC2, APOE and TTR in the ChP, the slides were imaged with a 63× oil-immersion objective using a confocal laser scanning microscope (Zeiss). Acquired images were then processed and analysed using ImageJ software.

### Quantitative histology

Hematoxylin eosin-stained ChP sections were examined for pathological changes by a board-certified pathologist with training and extensive experience in neurodegenerative disorders. Inflammatory cells, visualized either by hematoxylin eosin staining or by an anti-CD-68 antibody, were quantified on 6 high-magnification ChP images per section. Anti-CD-3 antibody recognized individual inflamatory cells on rare occasions in H and AD ChP. As a result, we did not pursue formal quantification of inflammatory cells recognized by the anti-CD-3 antibody. To measure NPC2, APOE and TTR total mean intensities, a grid of 500×500 pixel squares was placed over each ChP image. 30 grid squares were randomly selected in order to sample NPC2, APOE and TTR total mean intensities across the entire ChP. 3 rectangular regions of interest were examined per each grid square for a total of 90 regions of interest per image. The placement of the regions of interest was performed to best fit epithelium and stroma. Mean intensities of the regions of interest were calculated using Image Pro Premier 3D software (Media Cybernetics). To examine distribution of NCP2, APOE and TTR within the ChP, the slides were imaged with a 63× oil-immersion objective using a confocal laser scanning microscope (Zeiss). 1-4 ChP grape-like structures in 3 independent fields of view were sampled per each slide based on the ChP morphology for detailed analysis of the immunoreactivity of the choroid epithelium and stroma.

### Mass spectrometry proteomics of the CSF

CSF samples were analysed using two independent mass spectrometry proteomic protocols:

#### Protocol A

Samples were analyzed by LC-MS/MS using an UltiMate 3000 RSLCnano system (Thermo Fisher Scientific) coupled via an EASY-spray ion source (Thermo Fisher Scientific) to an Orbitrap Elite mass spectrometer (Thermo Fisher Scientific). Purified peptides were separated on 50 cm EASY-Spray column (75 µm ID, PepMap C18, 2 µm particles, 100 Å pore size; Thermo Fisher Scientific). For each LC-MS/MS analysis, 3 technical replicates of 500 ng of tryptic peptides per sample were used for 165 min runs. For the first 5 min, peptides were loaded onto a 2 cm trap column (Acclaim PepMap 100, 100 µm ID, C18, 5 µm particles, 100 Å pore size; Thermo Fisher Scientific) in loading buffer (98.9%/1%/0.1%, v/v/v, water/acetonitrile/formic acid) at a flow rate of 6 µl/min. Peptides were then added to buffer A (99.9%/0.1%, v/v, water/formic) and eluted from the EASY-Spray column with a linear 120 min gradient of 2% - 35% of buffer B (99.9%/0.1%, v/v, acetonitrile/formic acid), followed by a 5 min 90% B wash at a flow rate 300 nl/min. EASY-Spray column temperature was kept at 35°C. Mass spectrometry data were acquired with a Top12 data-dependent mode in the Orbitrap with a resolution of 120,000 at m/z 400, AGC 1E6 in the 300-1700 m/z range with a maximum injection time of 35 ms. The 2 m/z isolation window was used for MS/MS scans. Fragmentation of precursor ions was performed by CID with a normalized collision energy of 35. MS/MS data were acquired in ion trap with ion target value of 1E4 and maximum injection time of 100 ms. Dynamic exclusion was set to 70 s to avoid repeated sequencing of identical peptides.

#### Protocol B

Samples were analyzed by LC-MS/MS using an UltiMate 3000 RSLCnano system coupled to an Orbitrap Fusion Lumos tribrid mass spectrometer (both Thermo Fisher Scientific). 1 µg of tryptic peptides per sample (1 technical replicate per sample) was reconstituted in 5 µl of the solvent (98% water, 2% acetonitrile, 0.1% formic acid) and loaded on a 15 cm C18 Easy Spray column (2 µm particle size, 50 µm ID) kept at 45°C. Peptides were then separated using a binary solvent system (Solvent A: Water, 0.1% formic acid; Solvent B: Acetonitrile, 0.1% formic acid) operating at a flow rate of 300 nl/min with the following 120 min gradient: 2% to 19% B in 80 min, 19% to 30% B in 20 min, 30 to 98% B in 5 min, plateau at 98% for 2 min, return to initial conditions in 1 min and equilibration for 12 min. MS data were acquired using Xcalibur v4.1 (Thermo Fisher Scientific) in top-speed data dependent mode with MS1 in the Orbitrap (Resolution: 120,000, AGC: 2E5) followed by HCD (35% NCE) fragmentation of most abundant precursor ions with a charge state between 2-6 and detection in the Orbitrap (Resolution: 30,000, AGC: 5E4). Quadrupole isolation was enabled and dynamic exclusion was set to 60 s.

### CSF proteomic data processing

The raw data were analysed using MaxQuant software (version 1.5.6.5) by Andromeda search engine. Proteins were identified by searching MS and MS/MS data against the human proteome (UniProtKB, UP000005640, January 2017) and the common contaminants database. Carbamidomethylation of cysteines was set as a fixed modification. N-terminal acetylation and oxidation of methionines were set as variable modification. Trypsin was set as protease and maximum of two missed cleavages were allowed in the database search. Peptide identification was performed with an allowed initial precursor mass deviation up to 7 ppm (Orbitrap) and an allowed fragment mass deviation of 0.5 Da (ion trap). The “matching between runs” option was enabled to match identifications across samples within a time window od 20 s of the aligned retention times. The false discovery rate was set to 0.01 for both proteins and peptides with a minimum length of seven amino acids. LFQ was performed with a minimum ratio count of 2. Protein abundances were calculated on the basis of summed peptide intensities of “unique and razor” peptide.

For bioinformatic and statistical analysis, the MaxQuant output file (ProteinGroups.txt) was uploaded to the Perseus software (version 1.6.1.1). Proteins idenified by site and matching to the reverse database were excluded. Data was transformed to a logarithmic scale (log2(x)) and then normalized (Z-score). Samples were manually annotated based on technical and clinical conditions (AD, MCI, ALS, LYME). Technical replicates were averaged. Proteins were included into the analyses only if present in at least 50% of the samples in every condition. Differential expression was tested using *t*-test with permutation-based FDR calculation (p < 0.05). Volcano plot was used to visualize the results of the *t*-test. Clustvis 2.0 webtool was used for cluster analyses and Venn diagrams. g:Profiler was used for the functional enrichment analysis of the significant protein changes.

TMT-based quantitative discovery mode CSF preteomics dataset from a previously reported analysis of a well-established cohort of 297 individuals(Higginbotham et al., 2020; Johnson et al., 2020) was reanalyzed in this study for age-stratified differences (Fig. 2a-5, Fig. 3a).

### Processing of ChP tissue for mass spectrometry assays and global lipid analysis

ChP samples were homogenized using BeadBlaster 24 (Benchmark Scientific Inc.), lyophilized, resuspended in 100 µl of ultrapure water and the total protein concentrations determined using the BCA kit (Thermo Fisher Scientific). Next, 360 µl of isopropanol were added to each sample, which was vortexed, sonicated and centrifuged at room temperature. 400 µl of the supernatant were then removed and diluted 5-fold in 10% isopropanol for the ganglioside and other assays (lipid extract). Remaining pellets were dried at 37°C (Savant SDP121 P, SpeedVac, Thermo Fisher Scientific), reconstituted using glass beads in 100 µl of AmBic buffer (50 mM) with SDC (5 mg/ml), homogenized, vortexed and sonicated (protein extract). 20 µg of total protein per sample were next reduced (20 mM DTT; 10 min; 95 °C) and alkylated (40 mM IAA; 30 min; room temperature in the dark). Synthetic isotopically labeled standard peptides containing an enzymatically cleavable tag at C-terminus (TQL, JPT) were added to control for the variance in trypsin digestion. Trypsin was added to the samples in the 1:40 ratio (enzyme: total protein content, w/w), and the samples gently shaken overnight at 37°C. The digestion was quenched by adding 200 µl of 2% formic acid. Solid-phase extraction (SPE) was carried out on a mixed-mode cartridge according to the manufactureŕs instructions (Oasis Prime HLB).

### Validation of mass spectrometry targets

Gangliosides were quantified using selected reaction monitoring (SRM) assays in lipid extracts, which were internally standardized with 1.5 µM and 0.15 µM of isotopically labeled GM1 and GM3, respectively (Extended Data Figure 7). Proteins were also analyzed using SRM assays for proteotypic peptides as protein surrogates. Tentative identifications of proteotypic peptides detected in CP and concordant with the Skyline iRT Retention Time Prediction algorithm were validated using their isotopically-labeled analogs (Extended Data Figure 5). Only proteotypic peptides with the SRM signatures and retention times matching corresponding isotopically-labeled peptide standard were accepted as correct identifications. The peak area of the proteotypic peptide was normalized to the average peak area of 5 isotopically-labeled RT indices and the ratio multiplied by the concentration of RT indices to calculate relative protein concentrations using c(light) = peak area (light)/peak area (average heavy) x concentration (average heavy).

### Mass spectrometry quantitative protein assays

Dried solid-phase extracts of ChP digests were reconstituted in 13.3 µl of 5% acetonitrile with 0.1% formic acid. Samples were analyzed using ESI-UHPLC-SRM mass spectrometry on a triple quadrupole mass analyzer (1290 Infinity II and 6495B, Agilent) in positive ion detection mode. A volume of 2 µl was injected into the Peptide CSH C18 column (1.7 µm, 2.1 mm × 100 mm, Waters Corporation). The mobile phase at a flow rate of 0.3 ml/min consisted of buffer A (0.1% formic acid) and buffer B (0.1% formic acid in 95% ACN). Linear gradient elution was performed as follows: initial 5% B; 25 min 30 % B; 25.5 min 95 % B; 30 min 95 % B and from 31 to 35 min with 5 % B. The electrospray ionization source temperature was 200 °C and voltage 3.5 kV. SRM assays were scheduled for 2 min RT windows around peptide RT. SRM transitions (3-5 per proteotypic peptide) and optimal collision energies were obtained using the SRMAtlas database (www.srmatlas.org). Data were processed in Skyline (Version 19.1.0.193, MacCoss Lab).

### Mass spectrometry ganglioside assays

Gangliosides in lipid extracts of ChP were examined using both ESI-UHPLC-SRM positive and negative ion detection modes. Diluted lipid extracts were injected into a C18 column (CSH, 50 × 2.1 mm, 1.7 µm, Waters Corporation) thermostated at 40°C. The mobile phase of buffer A (0.5 mM ammonium fluoride) and buffer B (methanol: isopropanol (50:50 *v/v)* at the flow rate of 0.3 mL/min was used in positive ion mode. The gradient elution program (17.1 min) was: 0 min 30% B; 2 min 70% B ; 9 min 95 % B; 13 min 95% B; 13.3 min 5 % B; 14.3 min 5% B; 14.5 min 30% B; 17.1 min 30% B. The mobile phase of buffer A (0.5 mM ammonium fluoride and 10 mM ammonium acetate) and buffer B (acetonitrile: isopropanol (50:50; *v/v*) at a flow rate of 0.3 min mL/min was used in negative ion mode. The gradient elution program (19.1 min) was: 0 min 10% B; 4 min 85% B; 6.2 min 95% B; 10.2 95% B; 10.4 10% B; 14.4 95% B; 16.2 95 % B; 16.4 10% B; 19.1% B. The electrospray ionization source capillary voltage was 3500 V and 3000 V in positive and negative ion modes, respectively. The gas flow rate was 16 L/min and 14 L/min at 190°C, sheath gas pressure 20 PSI and 25 PSI at 350 °C and nozzle voltage 1300 V and 1500 V in positive and negative ion detection modes, respectively. Data were processed in Skyline (v19.1.0.193, MacCoss Lab). Gangliosides were quantified with the stable isotope-labeled GM3 using appropriate response factors (except for GM1 quantified with the stable isotope-labeled GM1). Ganglioside content was reported in ng/mg of CP tissue dry weight.

### Mass spectrometry lipid analysis

Orbitrap Fusion (Thermo Fisher Scientific) and UHPLC (Nexera X2, LC-30AD, Shimadzu) with identical C18 column (CSH, 50 × 2.1 mm, 1.7 µm, Waters Corporation) thermostated at 40°C and gradient elution program as specified for ganglioside assays in the positive ion mode were used to measure lipids. The electrospray ionization in positive and negative ion modes with capillary voltages of 4000 V and 3500 V, respectively, were used to acquire full-scan spectra. Sweep gas flow rate was 2 arbitrary units, sheath gas flow rate 30, auxiliary gas flow rate 5 and ion transfer tube and vaporizer temperatures 350 °C and 300 °C, respectively. The resolving power was 120 000 at 400 *m/z*, maximum injection time 5 ms, ACG 4.0×10^5^ and lenses RF level 50%. Data were acquired in centroid mode and converted to mzML format through MS Convert from ProteoWizard and analyzed through XCMS using the centWave algorithm. The XCMS output feature (*m/z*) list was submitted to the CEU mediator (Batch Advanced Search) to return database hits. The following CEU mediator settings were used: a) mass accuracy (tolerance) was set at 3 ppm, b) selected databases included LipidMaps, HMDB, Kegg and Metlin, c) respective ionization mode, d) input mass was *m/z* mass (instead of neutral mass) and e) in case of multiple hits per *m/z*, the tentative identification of adducts M+H+ was manually assigned based on similar retention time with a lipid from the same group, ppm error and available LipidMaps annotation. Features matching to multiple annotations with similar RT, ppm, and database availability remained unidentified. The most abundant lipid species were manually matched with the literature to verify tentative identifications further.

### Cholesterol spectrophotometric assay

Cholesterol was extracted from 1 mg of dry ChP tissue by adding 100 ul of Chloroform: Isopropanol. Samples were vortexed, sonicated and centrifuged. The ultra-filtrate was transferred to a separate vial and air-dried at 50°C to remove chloroform and placed in a vacuum concentrator centrifuge to remove remaining isopropanol. Cholesterol was quantified using a colorimetric assay based on the manufacturer’s instructions (MAK043, Sigma-Aldrich, USA).

### *In vivo* brain MRI imaging

T1-weighted 3-dimensional high-resolution MRI images were acquired using magnetization-prepared rapid gradient echo (MP-RAGE, TR/TE/TI = 2000/3.08/1100 ms, flip angle = 15°, 1.0 mm slice thickness) and fast spoiled gradient echo (FSPGR, TR/TE/TI = 7.26/2.99/400 ms, flip angle = 11°, 1.2 mm slice thickness) pulse sequences using 1.5 T (Siemens Avanto) and 3.0 T scanners (GE Signa HDxt) in the Czech Brain Aging Study and the Argentina Alzheimer’s Disease Neuroimaging Initiative, respectively. All images were first visually inspected to ensure appropriate data quality and to exclude participants with clinically relevant brain pathology that could interfere with cognitive functioning and testing such as subdural hematoma, cortical infarct, tumour or hydrocephalus. To correct for differences in head size, all volumetric measurements were normalized to the total intracranial volume.

### MRI intensity measurements

Based on FreeSurfer wrapped in R, all segmented regions of interest and their masks were exported, including ChP intensities. Since raw intensities of ChP preclude their direct use in statistical analysis, we normalized raw intensity of ChP to CSF considering MRI signal of the CSF does not significantly change between H and AD. The CSF strip algorithm was developed based on the white strip algorithm(Shinohara et al., 2014).

### MRI volumetry measurements

Measurements of cortical, hippocampal and ChP volumes were performed using FreeSurfer automated algorithm version v5.3 (http://surfer.nmr.mgh.harvard.edu/) as previously reported(Desikan et al., 2006; Fischl et al., 2002; Reuter et al., 2012). Subset of MRI brain images were further evaluated using 3D reconstructions. Automatic segmentation of FreeSurfer wrapped in R was used to obtain segmentation masks for all regions of interest. All T1-weighted images were registered to a common MNI305 template using linear (affine) and ANTs (SyN) spatial normalization algorithms(Avants et al., 2008) and symmetric image spatial normalization (SyN) algorithm with cross-correlation as similarity metric through Advanced Normalization Tools (ANTSs). Registered masks of structures of interest were then averaged to produce structure template used for visualizations. The iso-surfaces of the template were extracted using marching cube algorithm. Inverse warp field from SyN were then decomposed by Principal Component Analysis and the first principal component projected back to shape space and averaged across groups. Vector fields of differences between H and AD regions of interest were orthogonally projected to normal vector field from the average template, converted from mean to percentage change with normalization by centroid size and then used for coloring of the average template. To examine 3-dimensional differences between H and AD ChP in shape, ChP were represented with 4246 mesh points, the percentage change was calculated as (mean AD minus mean H) divided by mean H and multiplied by 100, where positive sign reflects larger areas in AD and scaling is done with respect to H mean.

### Manual MRI tracing

T1-weighted brain images underwent quality assessment, including dicom format check and check for imaging artefacts. ChP segmentation was performed manually by two trained readers with significant background and experience with MRI imaging. Segmentation was performed using Slicer (www.slicer.org) on a slice per slice basis with a paintbrush of 0.5 mm. Segmentation started at the top of the brain in the axial orientation with the reader moving down the brain to detect the first slice showing the contour of the ChP and then continuing consecutively through all the slices showing ChP. Once the region of interest was fully delineated, potential over- and under-segmented areas were checked in coronal orientation and segmentations corrected whenever necessary. Particular attention was given to brain regions where segmentation of the ChP may be difficult. First, ChP can be present in areas around the anterior pillars of the fornix and beneath there, it needs to be first distinguished from vascular structures and then segmented correctly. Second, signal from the hippocampal commissure and crura of the fornix, which appear as continuity of the ChP, was excluded from the segmentation. Particular attention was given to segment the thin lining of the ChP in the temporal horn of the lateral ventricles. After slice-to-slice manual segmentation of the ChP, the 3D volume rendering was performed to test for appropriate spatial appearance of ChP in line with the expected anatomy. Since ChP segmentations were performed voxel-wise, generating labelled maps, the volumes were calculated as the sum of all voxels of the labeled maps multiplied by voxel size. Considering small size of ChP the obtained volumes were not normalized.

## QUANTIFICATION AND STATISTICAL ANALYSIS

All statistical analyses were performed using commercially available softwares (R, Prism, Excel, SPSS and SAS). Mass spectrometry proteomics of the CSF and brain imaging analyses were performed using R (v.4.1.0, R Core Team 2020. R: A language and environment for statistical computing. R Foundation for Statistical Computing, Vienna, Austria). RColorBrewer, igraph, ggraph and ggplot2 packages were used for differential CSF protein expression analyses. For brain imaging analyses we used R packages Whitestripe, FreeSurfer, NeuroBase, fslr, fsbrain, misc3d, rgl and ggplot2. All values are expressed as mean ± s.e.m. Differences in means between H and AD were analysed using two-sample *t*-test at a significance α level of 0.05 unless otherwise specified. All tested hypotheses were two-sided (H_0_: equality of means vs H_1_: means are not equal). Missing values were not imputed. Differences in the shape of ChP between H and AD were assessed using a mesh point-by-point two-sample *t*-test with differences in coronal, sagittal and axial planes projected to H with *t*- statistics and p-values at a significance α level of 0.05 presented as a coloured statistical parameter map. No statistical analyses were performed to predetermine sample size.

## Data Availability

All the data are available upon request once the manuscipt is published.

https://www.synapse.org/#!Synapse:syn20835526

## ACKNOWLEDGMENTS

Authors wish to acknowledge Katja Klosterman and Sebastian Novotny and other members of the Stokin Lab, Michal Šitina and the Biostatistics core facility of the International Clinical Research Center of St. Anne’s University Hospital, Kateřina Coufalikova from the RECETOX Centre of the Faculty of Sciences, Masaryk University, Kathleen Myers of the Center for Neurologic Study and Jeffrey Metcalf and Sara Shuldberg from the Rissman Lab. We are grateful to the participants, families and all others involved in the Czech Brain Aging Study, Argentina Alzheimer’s Disease Neuroimaging Initiative, Shiley-Marcos ADRC at the UCSD, Imperial College Parkinson’s UK Brain Bank, Institute of Molecular and Translational Medicine, the ALS Tissue Bank and Target ALS Postmortem Core, and Barrow Neurological Institute. We thank PrecisionMed for their guidance regarding human tissues. This study was supported by the European Regional Development Funds No. CZ.02.1.01/0.0/0.0/16_019/0000868 ENOCH grant (S.B., J.D., G.F., J.F., M.H., J.H., S.K. and G.B.S.), Research Infrastructure RECETOX RI grant (No LM2018121) financed by the Ministry of Education, Youth and Sports, and Operational Programme Research, Development and Innovation - CZ.02.1.01/0.0/0.0/17_043/0009632 CETOCOEN EXCELLENCE project and the HORIZON 2020 No. 857560 (J.D., G.B.S. and Z.S.), the Czech Ministry of Health grants NV 18-04-00346 (Z.N.), NV 18-04-00455 and 00064203 (J.H.), NV19-08-00472 (Z.S. and K.S.), the No. CZ.02.1.01/0.0/0.0/16_026/0008451 INBIO grant (J.D.), NIH grant P30 AGO62429 (R.A.R.), the Barrow Neurological Foundation and the Fein Foundation (R.B.), the research infrastructure LM2018121 RECETOX grant (Z.S.), the Grant Agency of the Masaryk University No. MUNI/G/1131/2017 GAMU (Z.S.) and MUNI/A/1615/2020 (S.K.) grants, the National Cancer Institute of the NIH grant No. P30CA033572 (P.P.), the NIA U01 grant U01AG061357 (E.B.D. and N.T.S.) and the Institutional Support of Excellence 2. LF UK grant 6990332 (J.H.).

## AUTHOR CONTRIBUTIONS

G.B.S. designed and planned the study, M. Č. and G.B.S. analysed all of the data and wrote the manuscript, I.G.O. performed ChP cytokine experiments and analysis, C.L.-S. and R.A.R. provided ChP samples, C.L.-S. performed all the ChP neuropathological experiments and analysis, N.B., R.B., J.H., M.H. and K.S. contributed CSF samples, V.D.-D., K.G.-M., D.H., M.H., R.S., and P.P. performed all the CSF mass spectrometry experiments, J.D., D.J., M.N. and Z.S. performed all the ChP mass spectrometry and other lipid experiments, V.L. and K.T. performed ChP immunohistochemistry and analysis, H.C., J.H., Z.N., L.P. H.M., G.S. and M.V. were involved in the acquisition and analysis of MRI images, S.K. and M.V. performed MRI image and shape programming and analysis, including intensities and volumes, R.H. and T.V.V. and performed manual ChP measurements, M. Č., E.B.D. and D.H. performed all the bioinformatics assessments and analysis, S.B., J.D. and M. Š. performed biostatistical analysis, R.B., S.B., J.D., G.F., J.F., J.H., P.P., C.L.-S., N.T.S., G.S., Z.S. and R.A.S. provided continuous input during experiments and data analysis.

## DECLARATION OF INTERESTS

No competing interests.

## SUPPLEMENTAL INFORMATION TITLES AND LEGENDS

**Figure S1. No morphological changes in the ChP from H and AD**

(A) Representative image of haematoxylin eosin-stained ChP consisting of peripheral vasculature rich stroma lined by the cuboid epithelium.

(B) Percentage of ChP from healthy individuals (n=10) and patients with AD (n=10) showing epithelial atrophy, stromal fibrosis, vessel thickening, cystic dilations and calcifications. 1 technical replicate ± s.e.m.; two-sample *t*-test (p≤0.05).

(C) Average number of inflammatory cells in haematoxylin eosin-stained ChP from healthy individuals (n=10) and patients with AD (n=10) ChP. Mean number of inflammatory cells per 5 high power microscopy fields per section ± s.e.m.; two-sample *t*-test (p≤0.05).

(D) Representative low and high magnification images of ChP from healthy individuals and patients with AD stained with an anti-CD68 antibody (asterisks denote anti-CD68 antibody labelled cells in brown).

(E) Average number of anti-CD68 antibody labelled cells per ChP section in healthy individuals (n=8) and patients with AD (n=10) ChP. Mean number of inflammatory cells per 10 high power microscopy fields per section ± s.e.m.; two-sample *t*-test (p≤0.05).

(F) Representative low and high magnification image of a ChP from healthy individuals stained with an anti-CD3 antibody (asterisk denotes anti-CD3 antibody labelled cell in red).

**Figure S2. Lack of AD pathology in the ChP in AD**

Percentage of ChP from healthy individuals (n=8), patients with early (eAD, n=11) or late AD (lAD, n=13) and patients with AD and Parkinson’s disease (AD/PD, n=12) showing epithelial atrophy (A), stromal fibrosis (B), vessel thickening (C) and calcifications (D). 1 technical replicate ± s.e.m.; two-sample *t*-test (p≤0.05). Anti-amyloid-β antibody showing senile plaques in the hippocampus of patients with AD (positive control, E) but no senile plaques in the ChP from healthy individuals (F) or patients with eAD (G), lAD (H) or AD/PD (I) (magnification bar 100 µ). Anti-phospho-tau antibody showing neurofibrillary changes in the hippocampus of patients with AD (positive control, J), but no phospho-tau-immunoreactive deposits in the ChP from healthy individuals (K) or patients with eAD (L), lAD (M) or AD/PD (N) (magnification bar 100 µ). Anti-α-synuclein antibody reveals Lewy bodies in the temporal cortex of patients with AD (positive control, O), but no Lewy changes in the ChP from patients with H (P), eAD (R), lAD (S) or AD/PD (T) (magnification bar 100 µ).

**Figure S3. Unique and shared CSF proteins between AD and other disorders**

Venn diagram showing CSF proteins identified by both independent mass spectrometers and protocols in patients with mild cognitive impairment (MCI, n=10) and dementia (AD, n=22) due to AD, Lyme disease (Lyme, n=12) and amyotrophic lateral sclerosis (ALS, n= 14) or in any of the resulting combinations.

**Figure S4. CSF proteins changed in mild cognitive impairment, Lyme disease and amyotrophic lateral sclerosis compared with AD**

Volcano plots showing individual protein changes in the CSF of patients with MCI (n=10), Lyme disease (n=12) and ALS patients (n= 14) compared with patients with AD (n=22) using two independent mass spectrometers and protocols (A and B, p≤0.05).

**Figure S5. Interactions between CSF proteins changed in mild cognitive impairment, Lyme disease and amyotrophic lateral sclerosis compared with AD**

String plots depicting interactions between significantly increased and decreased CSF proteins in patients with MCI (n=10), Lyme (n=12) and ALS (n= 14) compared with patients with AD (n=22) using two independent mass spectrometers and protocols (A and B).

**Figure S6. Validation of the identity of the proteotypic peptides used in the ChP targeted protein mass spectrometry**

Skyline iRT retention time prediction algorithm shows identical chromatogram retention times between proteotypic peptides used to quantify ChP proteins (bottom) and stable isotopically labelled synthetic peptides (top). Peptides in A were used for relative quantification.

**Figure S7. No change in the cholesterol in the ChP between healthy individuals and patients with AD** Total and free cholesterol levels (ng/mg of dry ChP weight) in the ChP from healthy individuals (n=5) and patients with AD (n=5) ChP. 1 technical replicate ± s.e.m.; two-sample *t*-test (p≤0.05).

**Figure S8. Validation of the identity of the gangliosides used in the ChP targeted ganglioside mass spectrometry**

Identical retention times between proteotypic GM1 and GM3 and isotopically labeled GM1 and GM3. Gangliosides were quantified using selected reaction monitoring (SRM) assays internally standardized with 1.5 µM and 0.15 µM of isotopically labeled GM1 and GM3, respectively.

**Figure S9. No lipid changes in the ChP between healthy individuals and patients with AD**

No difference in the levels of phosphatidylcholines PC34:0, PC34:1, PC34:2, PC36:1 and PC36:2 (A), phosphatidylethanolamines PE-34:1, PE-36:2, PE-38:4 and PE-38:6 (B), sphingomyelins SM(34:1), SM(36:2) and SM(36:1) (C), phosphatidylinositols PI34:2, PI36:1, PI36:2, PI36:4, PI38:3, PI38:4 and PI38:5 (D), phosphatidylserines PS34:1, PS36:1, PS36:2 and PS38:4 (E), and sulfatides S36:1, S40:1, S41:1(OH), S41:2, S42:1, S42:1(OH), S42:2, S42:2(OH), S42:3, S43:2, S43:2(OH), S43:1(OH) and S44:2(OH) (F), in ChP between healthy individuals (n=5) and patients with AD (n=5). 6 technical replicates ± s.e.m.; two-sample *t*-test (p≤0.05).

**Figure S10. Increased ChP volumes in patients with AD compared with healthy individuals in a confirmatory cohort**

Normalized volumes of cerebellar cortices (negative control), hippocampi (positive control) and ChP in healthy individuals (n=13) and patients with AD (n=10). 1 technical replicate ± s.e.m.; two-sample *t*-test (p≤0.05).

## Supplementary information

Supplementary Figures S1-S10.

Supplementary Tables S1-3.

Supplementary Data S1-S11.

## Notes

### Competing Interest Statement

The authors have declared no competing interest.

### Author Declarations

IRB/ethical approvals were obtaine from: Czech Brain Aging Study, St. Anne's University Hospital, Brno, and Motol Hospital, Prague, Barrow Neurological Institute, Phoenix, Alzheimer's Disease Neuroimaging Initiative, Buenos Aires, Argentina, Shiley-Marcos Alzheimer's Disease Research Centre, University of California San Diego, Imperial College Parkinson's UK Brain Bank

