## Supplementary figures and images for "Pathogenesis of Alzheimer’s disease: Involvement of the Choroid Plexus"

### Figure S1

FIGURE S1

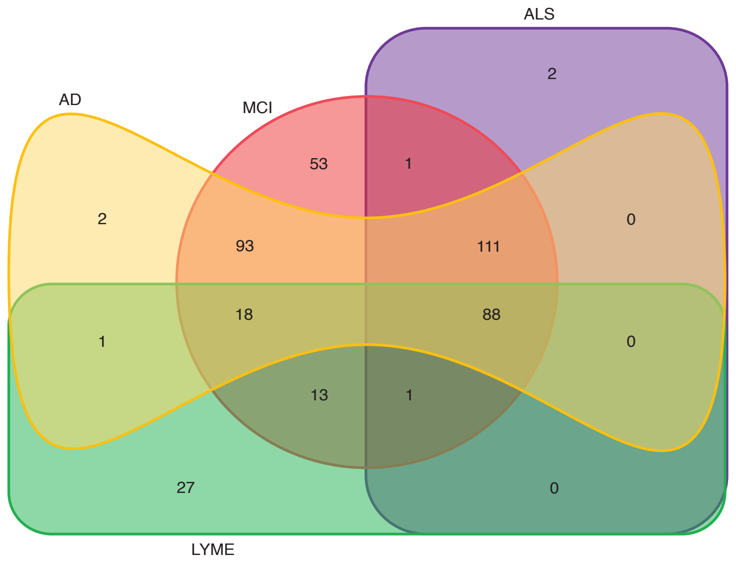

### Figure S2

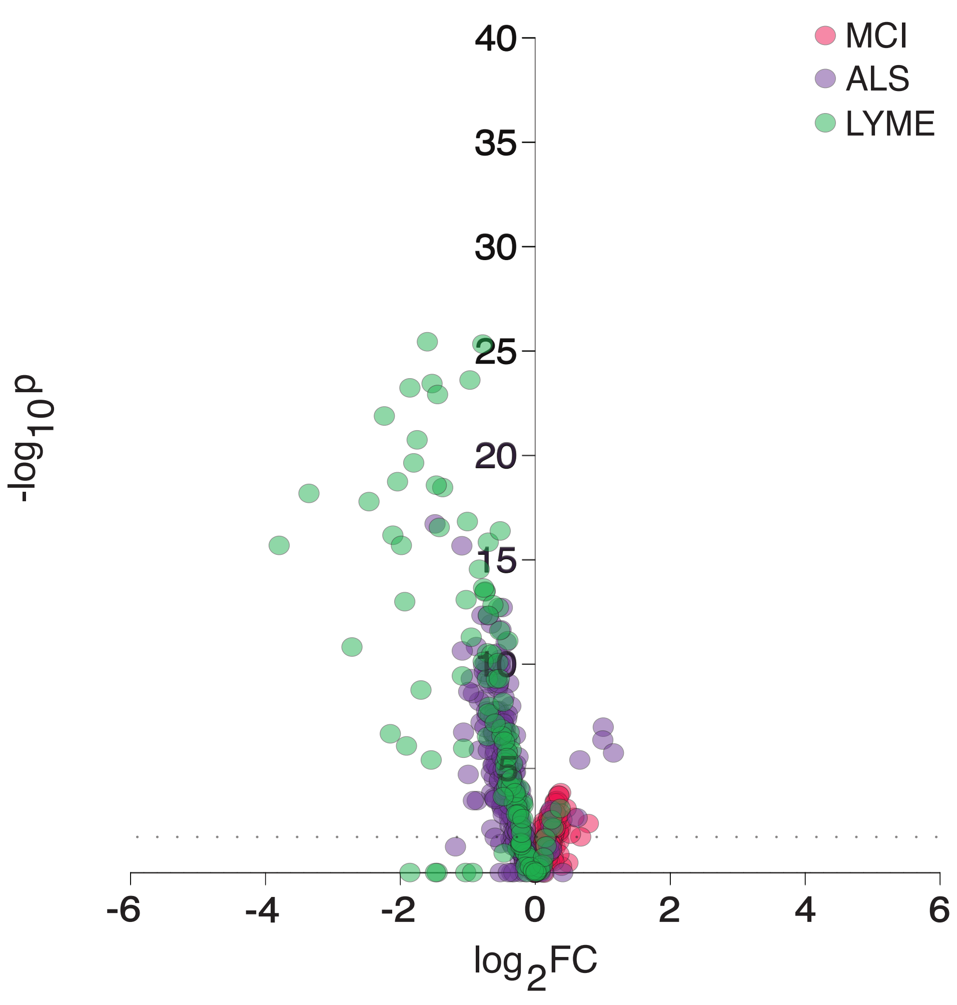

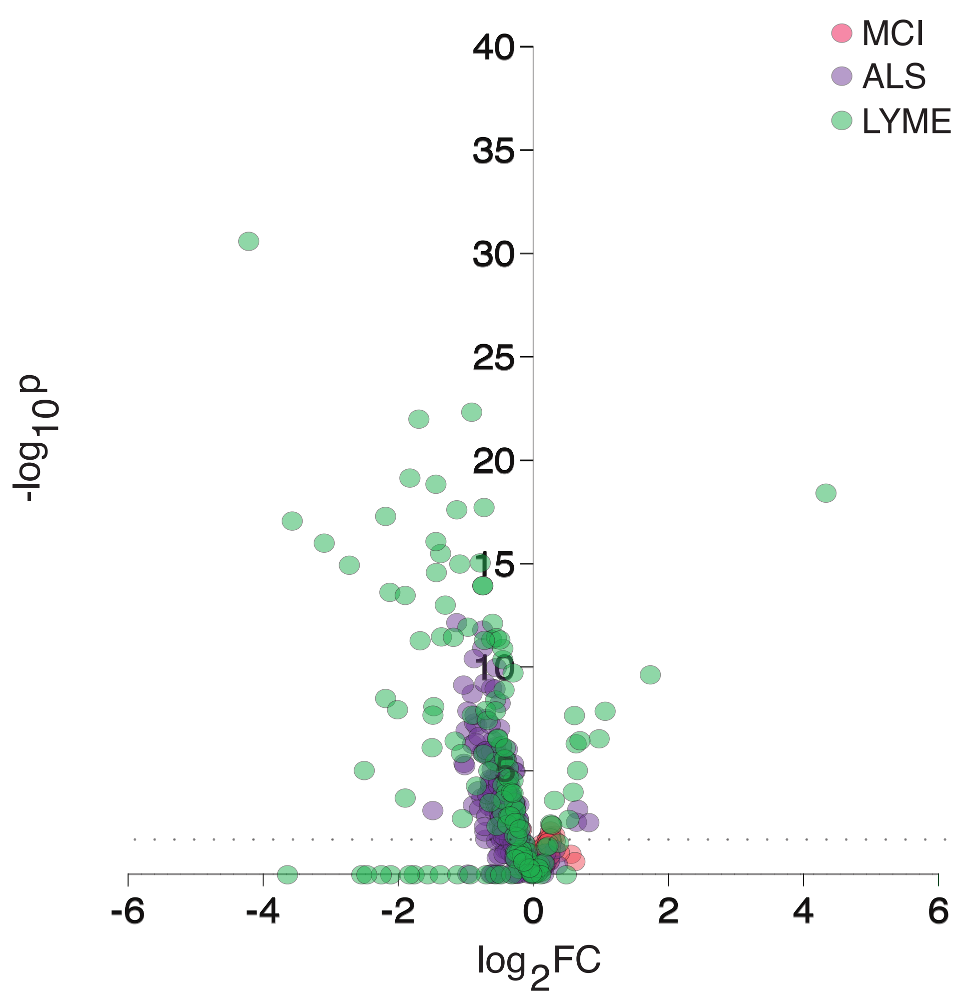

### Figure S4

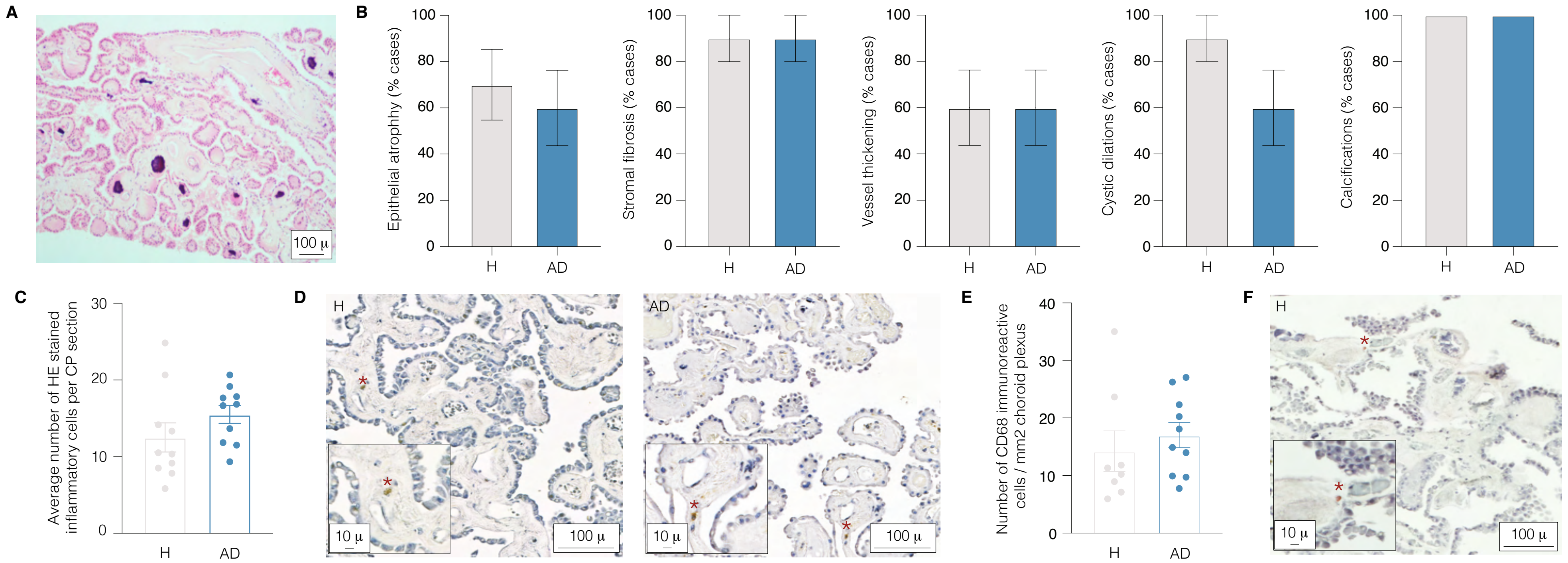

### Figure S5

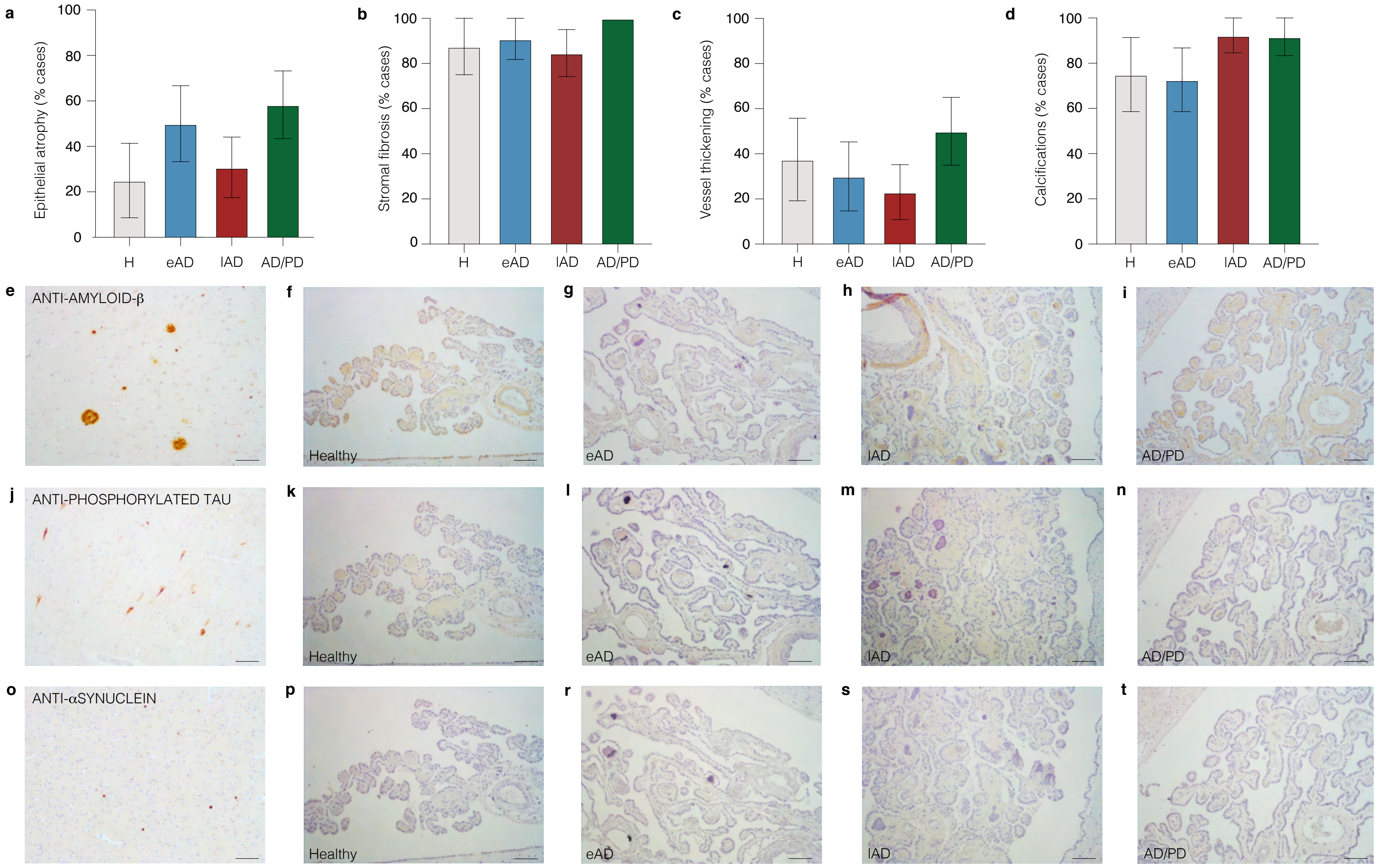

### Figure S6

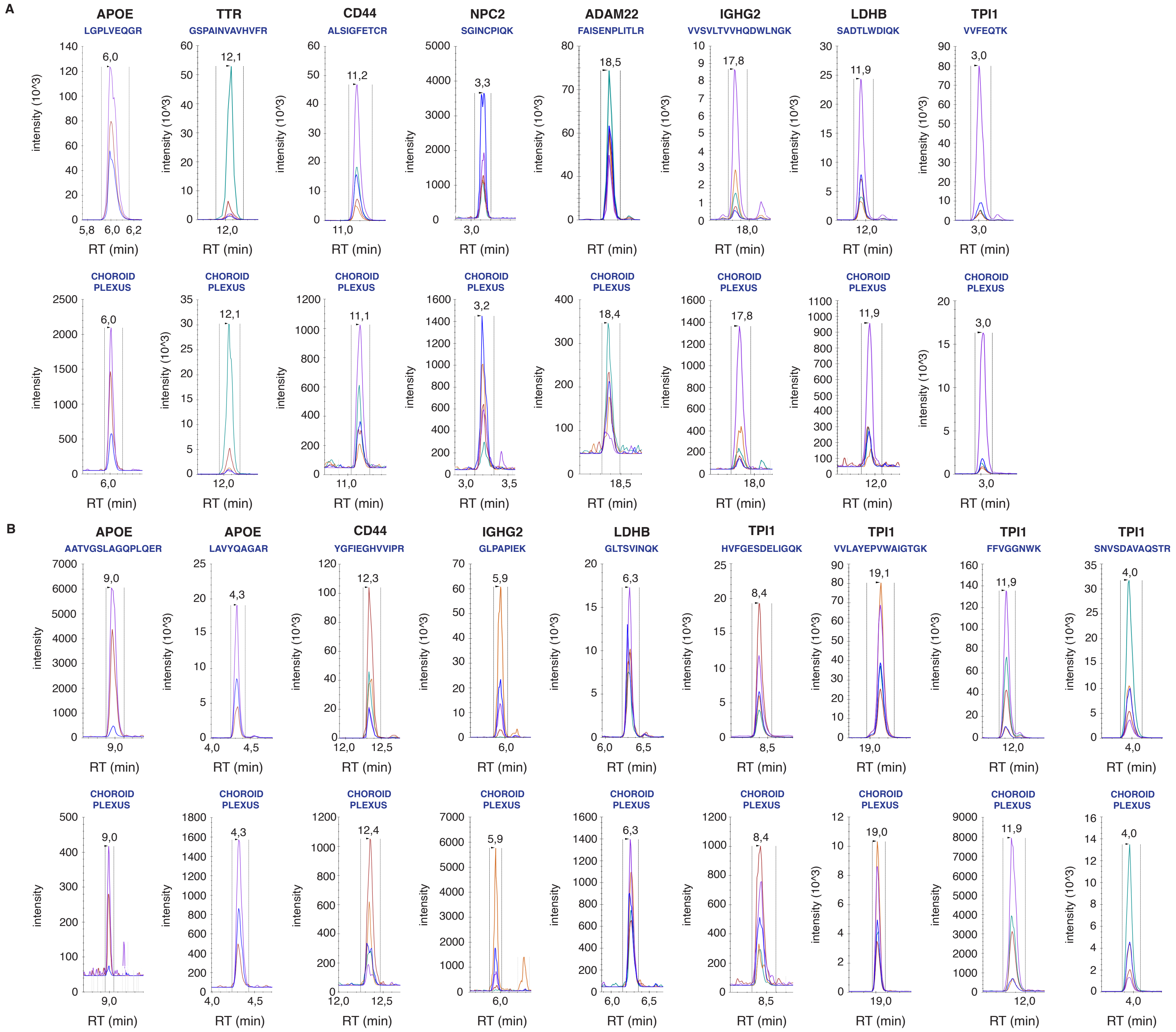

### Figure S7

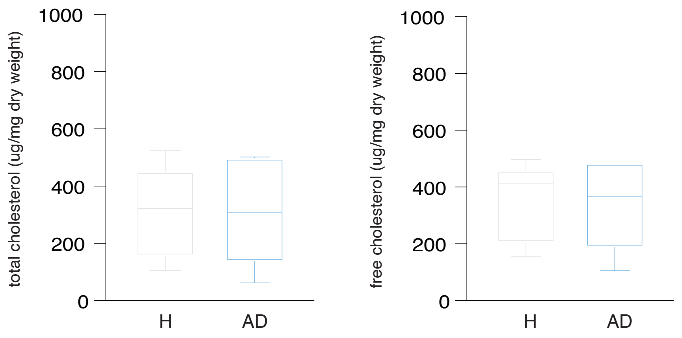

### Figure S8

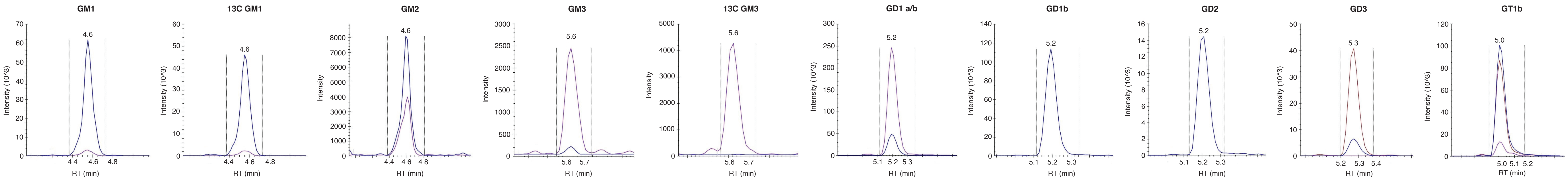

### Figure S9

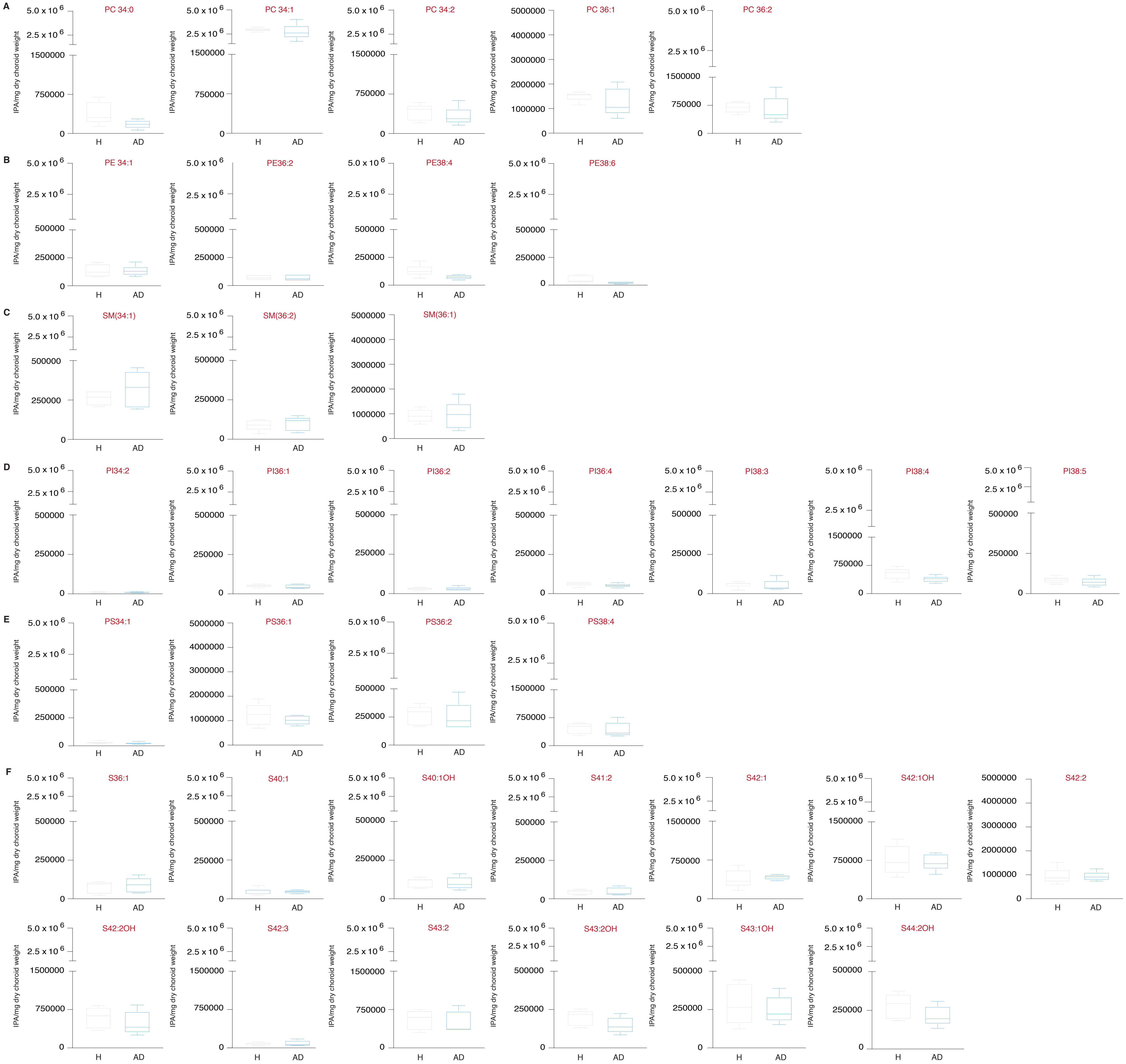

### Figure S10

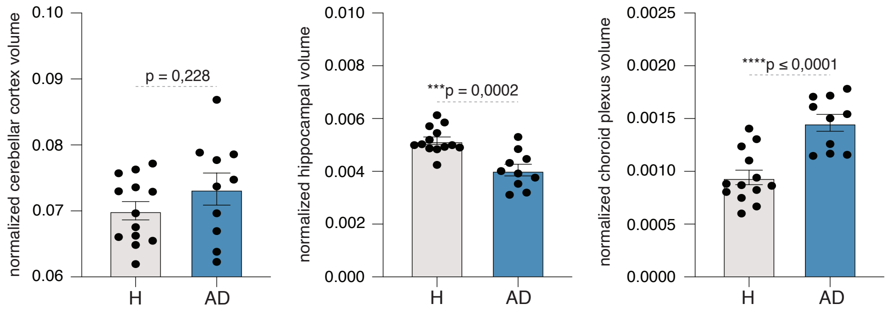
