## Supplementary material for "Pathogenesis of Alzheimer’s disease: Involvement of the Choroid Plexus": Figure S3

A

DECREASED IN MCI versus AD

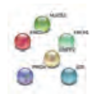

DECREASED IN ALS versus AD

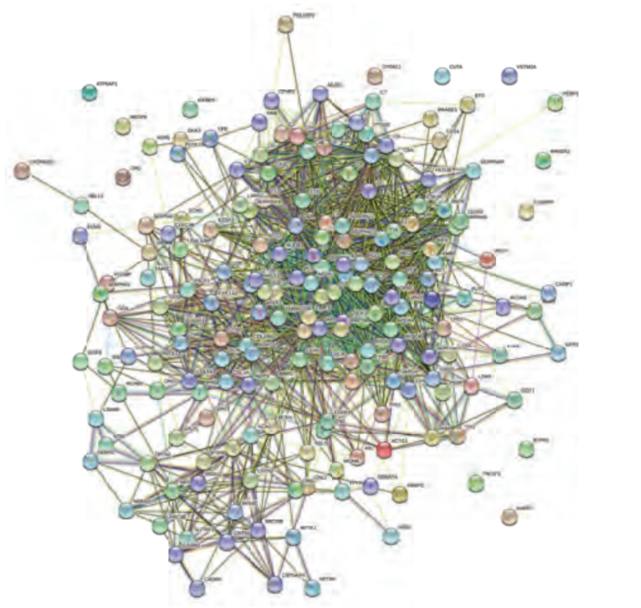

DECREASED IN LYME versus AD

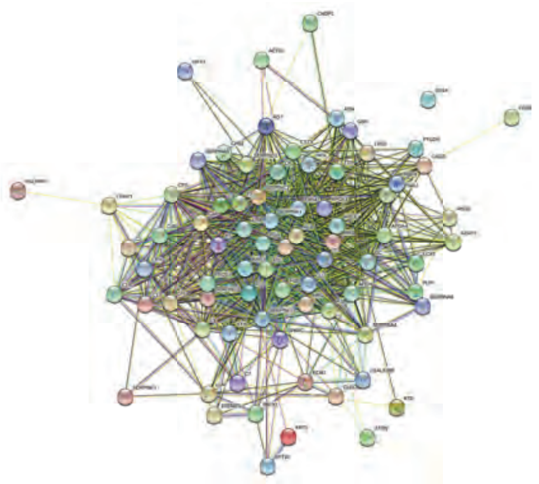

INCREASED IN MCI versus AD

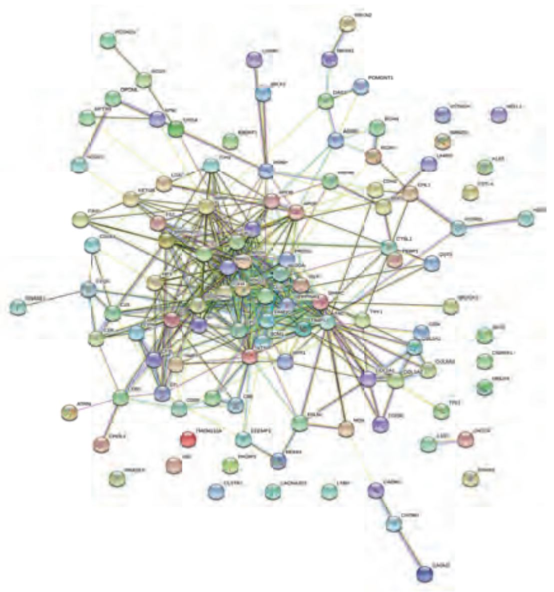

INCREASED IN ALS versus AD

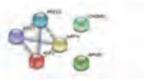

INCREASED IN LYME versus AD

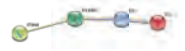

B

DECREASED IN MCI versus AD

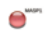

DECREASED IN ALS versus AD

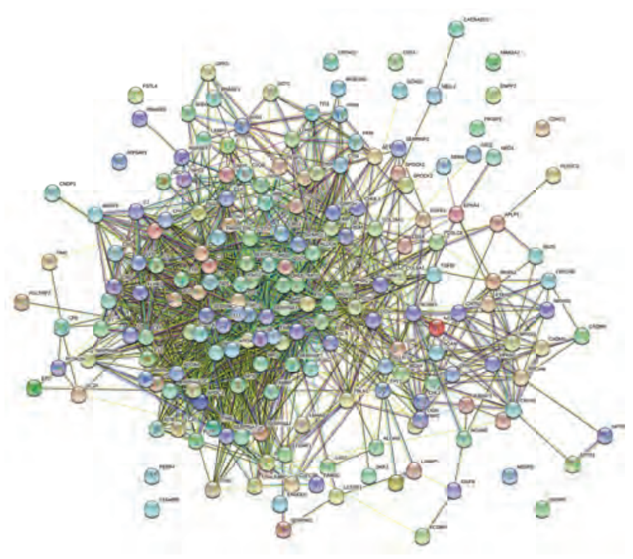

DECREASED IN LYME versus AD

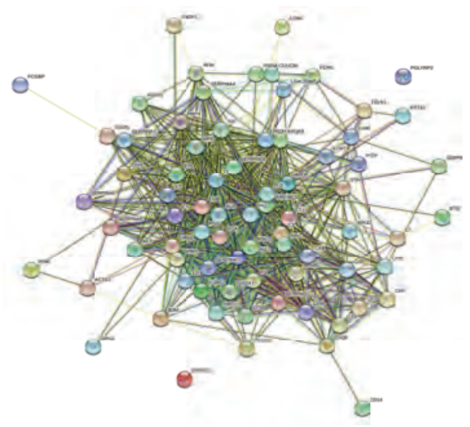

INCREASED IN MCI versus AD

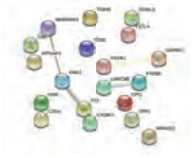

INCREASED IN ALS versus AD

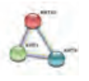

INCREASED IN LYME versus AD

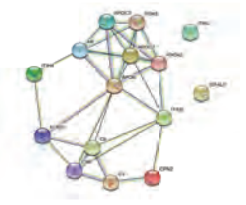
