## Supplementary material for "Pathogenesis of Alzheimer’s disease: Involvement of the Choroid Plexus": Data S1

| **Cohort characteristics** | **All subjects** | **H** | **AD** |
|---|---|---|---|
| # of subjects | 297 | 147 | 150 |
| Age (years±SD) | 66,62±6,36 | 65,05±8,16 | 68,16±8,29 |
| Sex (#) | | | |
| Male (0) | 111 | 41 | 70 |
| Female (1) | 186 | 106 | 80 |
| MoCA | 20,65±2,83 | 26,54±3,06 | 14,84±7,44 |
| CSF | | | |
| Aβ42 (pg/ml) | 422,47±194,40 | 546,76±159,42 | 299,85±107,71 |
| tTau (pg/ml) | 84,89±35,81 | 54,53±23,93 | 114,84±46,81 |
| pTau (pg/ml) | 47,59±20,56 | 32,15±14,43 | 62,82±24,00 |

Abbreviations:
H: Healthy
AD: Alzheimer's disease
MoCA: Montreal Cognitive Assessment

Source data:
Johnson E.C.B. et al (**2020**) *Nat Med* 26: 769-780.
cohort 1 (FNIH) at https://www.synapse.org/Consensus
Higginbotham L. et al (**2020**) *Sci Adv* 6: eaaz9360.
