## Supplementary material for "Pathogenesis of Alzheimer’s disease: Involvement of the Choroid Plexus": Data S2

**45-55 years old**

| **Cohort characteristics** | **All subjects** | **H** | **AD** |
|---|---|---|---|
| # of subjects | 29 | 19 | 10 |
| Age (years±SD) | 52,59±2,28 | 52,63±2,50 | 52,50±1,80 |
| Sex (#) | | | |
| Male (0) | 25 | 17 | 8 |
| Female (1) | 4 | 2 | 2 |
| MoCA | 20,48±10,04 | 25,47±6,43 | 11,00±8,75 |
| CSF | | | |
| Aβ42 (pg/ml) | 505,78±199,56 | 610,90±164,10 | 306,06±58,61 |
| tTau (pg/ml) | 86,32±60,68 | 54,44±24,46 | 146,88±62,77 |
| pTau (pg/ml) | 46,34±24,38 | 34,20±16,61 | 69,41±19,68 |

**56-65 years old**

| Cohort characteristics | All subjects | H | AD |
|---|---|---|---|
| # of subjects | 97 | 54 | 43 |
| Age (years±SD) | 60,44±3,00 | 60,44±2,98 | 60,44±3,02 |
| Sex (#) | | | |
| Male (0) | 59 | 38 | 21 |
| Female (1) | 38 | 16 | 22 |
| MoCA | 20,79±8,76 | 26,91±2,14 | 12,93±7,73 |
| CSF | | | |
| Aβ42 (pg/ml) | 437,18±203,10 | 557,84±177,97 | 282,05±103,83 |
| tTau (pg/ml) | 74,73±40,89 | 51,85±21,93 | 104,15±40,81 |
| pTau (pg/ml) | 44,59±24,43 | 33,04±14,75 | 59,45±26,32 |

**66-75 years old**

| **Cohort characteristics** | **All subjects** | **H** | **AD** |
|---|---|---|---|
| # of subjects | 133 | 64 | 69 |
| Age (years±SD) | 70,36±2,76 | 69,95±2,51 | 70,74±2,93 |
| Sex (#) | | | |
| Male (0) | 85 | 46 | 39 |
| Female (1) | 48 | 18 | 30 |
| MoCA | 21,21±7,32 | 26,67±1,97 | 16,14±6,80 |
| CSF | | | |
| Aβ42 (pg/ml) | 412,96±164,24 | 526,91±133,42 | 307,26±110,75 |
| tTau (pg/ml) | 88,29±50,63 | 55,08±23,01 | 119,09±49,79 |
| pTau (pg/ml) | 48,95±26,00 | 31,14±13,45 | 65,48±23,83 |

**76-90 years old**

| **Cohort characteristics** | **All subjects** | **H** | **AD** |
|---|---|---|---|
| # of subjects | 38 | 10 | 28 |
| Age (years±SD) | 80,00±3,87 | 82,10±4,35 | 79,25±3,38 |
| Sex (#) | | | |
| Male (0) | 17 | 5 | 12 |
| Female (1) | 21 | 5 | 16 |
| MoCA | 18,45±7,35 | 25,70±2,33 | 15,86±6,78 |
| CSF | | | |
| Aβ42 (pg/ml) | 355,04±147,74 | 492,11±144,69 | 306,09±114,18 |
| tTau (pg/ml) | 97,57±36,69 | 65,71±32,79 | 108,95±30,83 |
| pTau (pg/ml) | 51,32±22,38 | 29,98±12,60 | 58,95±20,06 |
