## Supplementary material for "Pathogenesis of Alzheimer’s disease: Involvement of the Choroid Plexus": Data S3

| Cohort characteristics | All subjects | **MCI** | **AD** | **ALS** | **Lyme** |
|---|---|---|---|---|---|
| # of subjects | 58 | 10 | 22 | 14 | 12 |
| Age (years±SD) | 67,3±10,0 | 73,2±7,0 | 71,6±9,0 | 55,8±8,7 | 69,7±4,3 |
| Sex (#) | | | | | |
| Male | 25 | 5 | 8 | 7 | 5 |
| Female | 33 | 5 | 14 | 7 | 7 |
| MMSE | 22,1±5,4 | 26,6±1,4 | 18,3±4,5 (12) | na | na |
| CSF | | | | | |
| Leukocytes/µ3 | 109,1±230,2 | 1,9±3,6 | 1,1±1,5 (13) | na | 334,2±300,9 (11) |
| Amyloid/Tau ratio | 0,8±0,4 | 0,9±0,4 | 0,7±0,3 (13) | na | na |

**Abbreviations:**
MCI: Mild cognitive impairment
AD: Alzheimer's disease
ALS: Amyotrophic lateral sclerosis
Lyme: Lyme disease
MMSE: Mini Mental State Examination

McKhann GM et al **2011** *Alzheimer's & Dementia* 7:263
Alberts MS et al **2011** *Alzheimer's & Dementia* 7: 270
Ludolph A et al **2015** *Amyotroph Lateral Scler Frontotemporal Degener* 16: 291
