## Supplementary material for "Pathogenesis of Alzheimer’s disease: Involvement of the Choroid Plexus": Data S4

| n | AGE (years±SD) | SEX (No F) | EDUCATION (years±SD) | APOE e4 (No subjects) | PATHOLOGICAL DIAGNOSIS | BRAAK (±SD) | MMSE (±SD) |
|---|---|---|---|---|---|---|---|
| 5 | 88±4,3 | 2 | 14,25±1,9 | 1 | Unremarkable | 2±0 | 26,4±3,6 |
| 5 | 88±4,3 | 3 | 15±3,3 | 3 | Alzheimer's disease | 5,8±0,4 | 19,8±5 |

Braak staging based on Braak H & Braak E **1991** *Acta Neuropathol* 82: 239

Abbreviations: No = number; F = females; MMSE = Mini Mental State Examination
