## Supplementary material for "Pathogenesis of Alzheimer’s disease: Involvement of the Choroid Plexus": Data S5

| n | AGE (years±SD) | SEX (No F) | EDUCATION (years±SD) | PATHOLOGICAL DIAGNOSIS | BRAAK (±SD) | MMSE (±SD) |
|---|---|---|---|---|---|---|
| 10 | 84,1±10,7 | 5 | 13,89±3,1 | Unremarkable | 1,32±0,8 | 28,3±2,8 |
| 10 | 84,1±10,4 | 5 | 15,7±2,8 | Alzheimer's disease | 5,7±0,8 | 15,9±8,5 |

Braak staging based on Braak H & Braak E **1991** *Acta Neuropathol* 82: 239
Abbreviations: No = number; F = females; MMSE = Mini Mental State Examination
