## Supplementary material for "Pathogenesis of Alzheimer’s disease: Involvement of the Choroid Plexus": Data S6

| Cohort characteristics | **H** | **early AD** | **late AD** | **AD & PD** |
|---|---|---|---|---|
| # of subjects | 8 | 11 | 13 | 12 |
| Age (years±SD) | 73,6±9,1 | 78,5±11,2 | 83,2±9,8 | 79,9±6,7 |
| Sex (#) | | | | |
| Male | 5 | 6 | 6 | 10 |
| Female | 3 | 5 | 7 | 2 |
| Neuropathology | | | | |
| Braak AD score | 0,6±0,9 | 2,6±0,7 | 4,5±0,8 | 4,3±1,2 |
| Braak PD score | 0±0 | 0,2±0,4 | 1,1±1,5 | 5,5±0,5 |
| CAA | 0±0 | 0,1±0,3 | 0,3±0,5 | 0±0 |
| Scoring hippocampi | | | | |
| Amyloid Plaques | 0±0 | 2,2±1,1 (9) | 2,4±1,0 | 1,8±1,3 |
| Neurofibrillary tangles | 0,8±0,8 (5) | 1,5±0,9 (10) | 2,3±0,5 | 2,1±0,8 |
| Lewy pathology | 0±0 | 0,8±1,2 (8) | 0,4±1,0 | 1,5±0,8 |

Hippocampi scoring criteria:

0 = absent
1 = scarce
2 = moderate
3 = abundant

Abbreviations:
H: Healthy
Early AD: Early Alzheimer's disease
Late AD: Late Alzheimer's disease
AD & PD: Alzheimer's and Parkinson's disease


Braak H & Braak E **1991** *Acta Neuropathol* 82: 239
Braak H et al **2006** *Acta Neuropathol* 112: 389
DicksonDW et al **2009** *Lancet Neurol* 8:1150
