## Supplementary material for "Pathogenesis of Alzheimer’s disease: Involvement of the Choroid Plexus": Data S7

| Cohort characteristics | All subjects | H | AD |
| --- | --- | --- | --- |
| # of subjects | 118 | 38 | 80 |
| Age (years±SD) | | | |
| | 73,0±7,6 | 70,0±6,4 | 75,0±7,6 |
| Sex (# of subjects) | | | |
| Male | 40 | 13 | 27 |
| Female | 78 | 25 | 53 |
| Education (years±SD, # of subjects) | | | |
| | 14,0±2,8 (104) | 16,0±2,3 (37) | 13,0±2,5 (67) |
| MMSE (Score±SD) | | | |
| | 23,0±4,5 (66) | 29,0±1,2 (15) | 21,0±3,2 (51) |
| *APOE* status (# of subjects, %) | | | |
| *E4/E4* | 7 (8,6%) | 0 (0%) | 7 (12,7%) |
| *E4/E3* | 31 (38,3%) | 3 (11,5%) | 28 (50,9%) |
| *E3/E3* | 35 (43,2%) | 17 (65,4%) | 18 (32,7%) |
| *E3/E2* | 8 (9,9%) | 6 (23,1%) | 2 (3,6%) |
| *E2/E2* | 0 (0%) | 0 (0%) | 0 (0%) |
