## Supplementary material for "Pathogenesis of Alzheimer’s disease: Involvement of the Choroid Plexus": Data S8

| Cohort characteristics | All subjects | H |
|---|---|---|
| # of subjects | 59 | 23 |
| Age (years±SD) | | |
| | 73,4±7,4 | 71,3±4,9 |
| Sex (# of subjects) | | |
| Male | 19 | 8 |
| Female | 40 | 15 |
| Educaton (years±SD, # of subjects) | | |
| | 14,1±2,5 (58) | 15,6±2,2 (22) |
| MMSE (Score±SD) | | |
| | 24,2±5,0 (57) | 29,2±1,5 (23) |
| *APOE* status (# of subjects, %) | | |
| *E4/E4* | 3 (7,9%) | 0 (0%) |
| *E4/E3* | 15 (39,5%) | 2 (13,3%) |
| *E3/E3* | 14 (36,8%) | 8 (53,3%) |
| *E3/E2* | 6 (15,8%) | 5 (33,3%) |
| *E2/E2* | 0 (0%) | 0 (0%) |

| AD |
| --- |
| 36 |
| 74,6±8,3 |
| 11 |
| 25 |
| 13,2±2,2 (36) |
| 20,6±3,3 (34) |
| 3 (13,0%) |
| 13 (56,5%) |
| 6 (26,1%) |
| 1 (4,4%) |
| 0 (0%) |
