## Supplementary material for "Pathogenesis of Alzheimer’s disease: Involvement of the Choroid Plexus": Data S9

| Cohort characteristics | All subjects | H | AD |
|---|---|---|---|
| # of subjects | 9 | 4 | 5 |
| Age (years±SD) | | | |
| | 75,0±5,0 | 75,0±5,7 | 76,0±4,6 |
| Sex (# of subjects) | | | |
| Male | 4 | 2 | 2 |
| Female | 5 | 2 | 3 |
| Education (years±SD, # of subjects) | | | |
| | 14,0±2,0 (9) | 16,0±1,8 (4) | 13,0±1,4 (5) |
| MMSE (Score±SD) | | | |
| | 23,0±5,9 (9) | 29,0±0,9 (4) | 19,0±3,2 (5) |
| *APOE* status (# of subjects, %) | | | |
| *E4/E4* | 1 (16,7%) | 0 (0%) | 1 (25,0%) |
| *E4/E3* | 4 (66,7%) | 1 (50,0%) | 3 (75,0%) |
| *E3/E3* | 0 (0%) | 0 (0,0%) | 0 (0%) |
| *E3/E2* | 1 (16,7%) | 1 (50,0%) | 0 (0%) |
| *E2/E2* | 0 (0%) | 0 (0%) | 0 (0%) |
