## Supplementary material for "Pathogenesis of Alzheimer’s disease: Involvement of the Choroid Plexus": Data S10

**Data S11**

| Cohort characteristics | All subjects | H | AD |
|---|---|---|---|
| # of subjects | 19 | 9 | 10 |
| Age (years±SD) | | | |
| | 71,0±6,4 | 72,0±4,8 | 70,0±7,7 |
| Sex (# of subjects) | | | |
| Male | 7 | 4 | 3 |
| Female | 12 | 5 | 7 |
| Education (years±SD, # of subjects) | | | |
| | 15,0±2,3 | 16,0±1,9 | 14,0±2,2 |
| MMSE (Score±SD) | | | |
| | 25,0±4,5 | 29,0±1,7 | 22,0±3,3 |
| *APOE* status (# of subjects, %) | | | |
| *E4/E4* | 2 (15,4%) | 0 (0%) | 2 (28,6%) |
| *E4/E3* | 5 (38,5%) | 1 (16,7%) | 4 (57,1%) |
| *E3/E3* | 3 (23,1%) | 2 (33,3%) | 1 (14,3%) |
| *E3/E2* | 3 (23,1%) | 3 (50,0%) | 0 (0%) |
| *E2/E2* | 0 (0%) | 0 (0%) | 0 (0%) |
