## Supplementary material for "Pathogenesis of Alzheimer’s disease: Involvement of the Choroid Plexus": Data S11

**Data S10**

| Cohort characteristics | All subjects | H | AD |
|---|---|---|---|
| # of subjects | 23 | 13 | 10 |
| Age (years±SD) | 71,0±7,0 | 68,1±7,0 | 74,7±5,1 |
| Sex (#) | | | |
| Male | 8 | 4 | 4 |
| Female | 15 | 9 | 6 |
| Cognitive status | | | |
| MMSE (score±SD) | 24,5±5,8 | 28,7±1,4 | 19,1±4,8 |
