## Supplementary material for "Pathogenesis of Alzheimer’s disease: Involvement of the Choroid Plexus": Table S1

Table S1 – List of significantly changed CSF proteins in AD versus H split by age groups.

| | |
|---|---|
| 45-55 *vs* 56-65 *vs* 66-75 *vs* 76-90 | NPTX2, PKM, ALDOA, LDHA, SPP1, ENO2, YWHAZ |
| 45-55 *vs* 56-65 *vs* 66-75 | GAP43, GOT1, LDHB, PARK7, BASP1, PEBP1, CHI3L1, MDH1, PGK1 |
| 45-55 *vs* 66-75 *vs* 76-90 | SPARC |
| 56-65 *vs* 66-75 *vs* 76-90 | P02649-4, SMOC1 |
| 45-55 *vs* 56-65 | ENO1, PPIA, PTPRN2 |
| 45-55 *vs* 66-75 | CD163, ALCAM, GDA, CALM2, GMFB, APOC3, KN45-55, FN1, SOD1, TRPC1 |
| 45-55 *vs* 76-90 | LMAN2, LTBP1 |
| 56-65 vs 66-75 | FGG, GAPDH, CAMK2B, SPON1, ZP2, FGB, ALDOC, C1QTNF3, NPY |
| 56-65 *vs* 76-90 | VSTM2B, B4GAT1, ECM1 |
| 45-55 | VSI76-90, MCAM, WFIKKN2, NA, MGAT1, CBLN3, VCAN, PRELP, FETUB, CACNA2D1, FAT2, MRC1, ADAMTS4, MFGE8, SELENBP1, HTRA1, NCAM2, QDPR |
| 56-65 | FGA, SC66-75, CDH6, CNTN1, PAM, MARCKS, PCSK1, NELL2, F5, IL6ST, KIT, NRCAM, CANX, CELSR2, NRXN3, NPTX1, CHGA, ST6GAL2, CD99, NUCB2, CD55, PLXDC2, RNASE4, NPTXR, SC56-65, DDAH2, NPPC, VGF |
| 66-75 | PON1, TXN, IGHV3-74, C4BPA, PLG, B4E1Z4, SERPINC1, BCHE, DYNLL2, GC, LUM, AZGP1, ANG, ITIH5, HRG, DNER, SOD2, UBA52, EFEMP2, SERPINA6, CA1, SERPINF2, ITIH2, PTGDS, PRDX1, NCAM1, AMBP, RBP4, C2, C2orf40, GM2A, SEZ6, NID2, FRZB, ITIH4, PRNP, COL1A1, APOA2, APOC1, PAPLN, ORM2, CFI, GPX3, SEZ6L2, SCR45-55, HBD, VTN, NPC2, CTSA, ATRN, HBA2, APOA4, KLKB1, SHISA5, F12, CD14 |
| 76-90 | SOD3, CFHR2, DCN, PRKCSH, CTSZ, OMD, ENDOD1, PTPRS, TGFBR3, CDH13, FCGBP, ART3, MCFD2, TIMP2, NTN45-55, FSTL4 |
