## Supplementary material for "Pathogenesis of Alzheimer’s disease: Involvement of the Choroid Plexus": Table S2

Table S2 – List of unique and shared CSF proteins between MCI, AD, ALS and Lyme identified using mass spectrometry protocol A and B.

| | |
|---|---|
| MCI & AD & ALS & LYME | ITIH1, C4B, PLG, PGLYRP2, GC, LUM, C1S, C1R, IGHM, FBLN1, C8A, HRG, GSN, C5, SERPINA6, P07225, TTR, P01780, SERPINF1, LGALS3BP, F5, AMBP, CNDP1, RBP4, IGHG4, CFB, LRG1, A2M, SERPINA7, F2, ORM2, CFI, AHSG, GPX3, IGKC, HPX, VTN, IGLL5;IGLC1, APOE, APOA4, C3, F12, APOH, FGA, ORM1, PON1, ALB, IGHG2, SERPINC1, SERPING1, CON__P15636, APOA1, AZGP1, CLU, AFM, TF, APOD, SERPINA3, A1BG, IGHA1, SERPINF2, APOB, CFH, C7, ITIH2, AGT, P0DOY3;P0DOY2, C2, C4A, CST3, CON__P00761, IGHG1, CP, SERPIND1, FCGBP, CLEC3B, ITIH4, SERPINA1, C1QB, SERPINA4, IGHG3, C6, KNG1, FN1, C9, KLKB1, CD14, HP |
| MCI & AD & ALS | MMP2, SCG3, WFIKKN2, CHGB, RNASE1, CADM4, FAM3C, CD163, PAM, KLK6, CLSTN1, KRT9, CHL1, NTM, CACNA2D1, GOT1, PTPRD, OGN, PKM, PCOLCE, CDH13, NFASC, PSAP, PTGDS, BCAN, THY1, NRCAM, PTPRZ1, NRXN3, NPTX1, ATP6AP1, GM2A, KRT1, SPP1, ISLR, DKK3, CRTAC1, COL1A1, ALDOC, KRT10, CADM3, B2M, SCG2, NOV, SERPINI1, NEGR1, MEGF8, SOD1, NPC2, NCAM2, CPE, PLTP, VGF, IGFBP2, TPI1, BTD, COL6A1, SEMA7A, IGFBP6, NCAN, NEO1, SPARC, MCAM, SOD3, FGG, CSF1, CNTN1, OPCML, UBA52;RPS27A;UBB;UBC, ALCAM, CTSD, ENPP2, RELN, LRRC4B, NRXN1, LDHB, FAT2, SIRPA;SIRPB1, PENK, DAG1, TGFBI, TIMP1, NELL2, ACTG1, IGFBP7, SPARCL1, FGB, CDH2, NCAM1, SCG5, CNTN2, PEBP1, CHI3L1, CHGA, EPHA4, B4GAT1, PRNP, CFD, NPTXR, APP, LSAMP, ECM1, APLP1, EFEMP1, PCSK1N, IGSF8, CADM1, SPOCK1, KRT2, PIK3IP1, FSTL4 |
| MCI & AD & LYME | C1QC, IGHV3-72, APOA2, CFHR1, C8B, APOC3, LCAT, IGHA2, C8G, CPN2, IGFALS, IGKV2D-28, IGKV4-1, APOC2, HBB, CPB2, ATRN, HBA1 |
| MCI & ALS & LYME | IGHV4-61 |
| MCI & AD | IDS, ENO1, PRCP, PTN, CGREF1, SEZ6L, GGH, OMD, MAN1A1, CTSB, LMAN2, NID1, |

| | |
|---|---|
| | MFAP4, FETUB, PEBP4, ITIH5, PTPRS, ECM2, FGFR1, AXL, LAMP2, CD59, RNASET2, CALR, PRG4, RNASE6, PTPRN2, LYZ, LDHA, QSOX1, AGRN, COL6A3, CD55, MAN2A2, PLXDC2, ENO2, IMPAD1, LTBP4, MASP1, TPP1, PTPRF, CNTNAP4, MDH1, TIMP2, POMGNT1, LGALS1, PCMT1, VSIG4, HSPA5, DCN, PRELP, SPOCK2, PPIA, MINPP1, CADM2, ICOSLG, COL18A1, ENDOD1, GAPDH, FBLN2, C16orf89, QPCT, NUCB1, CSF1R, PTPRG, GDI1, TCN2, TMEM132A, APLP2, CNTFR, CTSL, IL6ST, OMG, CPQ, ALDOA, HSPG2, CUTA, COL1A2, SERPINA5, SEZ6, EXTL2, SELENBP1, LY6H, GSTP1, SCRG1, LPHN3, NRXN2, VASN, VSTM2A, FBLN5, MEGF10, YWHAZ, PGK1 |
| AD & LYME | P0DP03;P01768 |
| MCI & ALS | IGF2 |
| MCI & LYME | P04432;P01597, P80748, SAA4, IGHV5-51, P01714, SEPP1, P01701, C4BPA, P01619, IGKV2-40, IGKV1-5, APOM, APOC1 |
| AD | SLITRK1, CON__P02769 |
| MCI | NPDC1, L1CAM, ATP6AP2, COL3A1, PVRL1, RGMB, VCAM1, NUTF2, SUSD5, SEMA3G, SOD2, VSTM2B, GDA, MMRN2, OLFML3, MFGE8, PLD3, ART3, RARRES2, GPR37L1, LRP1, ADAMTS1, PAPLN, SEZ6L2, CD99L2, LPHN1, IGKV1-8;IGKV1-9, SGCE, LYVE1, PI16, FMOD, C1QA, MAN2A1, GRIA4, PPIB, CLSTN3, CAMK2A, PVALB, COMP, EFNB2, CXCL16, TNFRSF21, TYRO3, HEXA, MIA, CNTNAP2, CD99, ICAM5, RNASE4, PLXNB2, CANT1, PCDHGC5, IGKV2-24 |
| ALS | KRT14, KRT6C |
| LYME | HPR, P06310, P01782;P0DP04, IGKV3D-11, CPN1, LPA, CD5L, VWF, ITIH3, IGKV1-27, P01717, F13B, LBP, P01706, APOL1, THBS1, P23083, PPBP, PZP, APCS, HABP2, P01624, IGJ, SAA1, APOF, P01599, GPLD1 |
