## Supplementary material for "Pathogenesis of Alzheimer’s disease: Involvement of the Choroid Plexus": Table S3

Table S3 – List of unique and shared CSF proteins between MCI, ALS and Lyme compared with AD identified by two independent mass spectrometry protocols.

| | |
|---|---|
| MCI & ALS & LYME | ITIH1, C4B, PLG, SEZ6L, RNASE1, PGLYRP2, CD163, GC, LUM, C1S, C1R, IGHM, KRT9, FBLN1, FETUB, C1QC, C8A, HRG, IGHV3-72, GSN, C5, SERPINA6, P07225, PKM, CDH13, TTR, P01780, SERPINF1, LGALS3BP, LYZ, F5, PTGDS, AMBP, NRCAM, CNDP1, RBP4, IGHG4, LRG1, CFB, QSOX1, A2M, KRT1, SERPINA7, DKK3, CRTAC1, F2, MAN2A2, APOA2, KRT10, ORM2, CFI, AHSG, B2M, CFHR1, GPX3, IGKC, HPX, MASP1, C8B, APOC3, VTN, PLTP, IGLL5;IGLC1, BTD, LCAT, IGHA2, APOE, APOA4, C3, F12, APOH, FGA, ORM1, PON1, SPARC, FGG, ALB, IGHG2, SERPINC1, C8G, CNTN1, SERPING1, CON__P15636, APOA1, AZGP1, CLU, AFM, TF, APOD, RELN, SERPINA3, A1BG, CPN2, IGFALS, SERPINF2, IGHA1, IGKV2D-28, TGFBI, APOB, CFH, C7, ITIH2, ACTG1, FGB, IGKV4-1, AGT, P0DOY3;P0DOY2, C2, C4A, CST3, CON__P00761, PEBP1, IGHG1, SERPINA5, CP, SERPIND1, FCGBP, CLEC3B, ITIH4, SERPINA1, C1QB, CFD, SERPINA4, APOC2, IGHG3, CON__P02769, ECM1, C6, HBB, KNG1, FN1, C9, CPB2, ATRN, EFEMP1, KLKB1, HBA1, P0DP03;P01768, CD14, HP, FSTL4 |
| MCI & ALS | SCG3, MMP2, IDS, WFIKKN2, ENO1, PRCP, PTN, CGREF1, CHGB, CADM4, GGH, FAM3C, PAM, KLK6, OMD, CLSTN1, CTSB, NID1, LMAN2, PEBP4, ITIH5, CHL1, NTM, CACNA2D1, PTPRS, ECM2, FGFR1, GOT1, AXL, LAMP2, PTPRD, CD59, OGN, RNASET2, PCOLCE, CALR, RNASE6, NFASC, PTPRN2, PSAP, THY1, BCAN, LDHA, PTPRZ1, NRXN3, NPTX1, ATP6AP1, AGRN, GM2A, COL6A3, SPP1, ISLR, COL1A1, CD55, ALDOC, PLXDC2, CADM3, ENO2, SCG2, NOV, SERPINI1, NEGR1, MEGF8, LTBP4, TPP1, CNTNAP4, MDH1, TIMP2, SOD1, NPC2, NCAM2, CPE, POMGNT1, VGF, IGFBP2, TPI1, PCMT1, COL6A1, SEMA7A, IGFBP6, NCAN, NEO1, MCAM, SOD3, HSPA5, CSF1, DCN, PRELP, SPOCK2, PPIA, OPCML, MINPP1, UBA52;RPS27A;UBB;UBC, CADM2, ALCAM, |

| | |
|---|---|
| | ICOSLG, COL18A1, ENDOD1, GAPDH, CTSD, ENPP2, C16orf89, LRRC4B, QPCT, NRXN1, NUCB1, LDHB, CSF1R, FAT2, PTPRG, SIRPA;SIRPB1, GDI1, PENK, DAG1, TMEM132A, TIMP1, APLP2, NELL2, IGFBP7, SPARCL1, CDH2, CTSL, NCAM1, IL6ST, OMG, CPQ, SCG5, ALDOA, HSPG2, CUTA, CNTN2, COL1A2, CHI3L1, SEZ6, CHGA, EXTL2, SELENBP1, EPHA4, B4GAT1, PRNP, LY6H, GSTP1, NPTXR, SCRG1, APP, LPHN3, LSAMP, APLP1, NRXN2, VASN, VSTM2A, FBLN5, PCSK1N, IGSF8, CADM1, MEGF10, SPOCK1, KRT2, PIK3IP1 |
| MCI & LYME | MFAP4, PRG4 |
| MCI | MAN1A1, IMPAD1, PTPRF, LGALS1, VSIG4, FBLN2, TCN2, SLITRK4, CNTFR, YWHAZ, PGK1 |
| ALS | SLITRK1 |
| LYME | IGKV3D-11 |
